# Clustering of Multimorbidity and Social Care Needs: 10-year all-cause mortality in a cohort study of more than 7 million people

**DOI:** 10.1101/2025.05.21.25328077

**Authors:** Nazrul Islam, Alim Sabary, Lucy Smith, Tassella Isaac, Glenn Simpson, Miriam Santer, Andrew Farmer, Hajira Dambha-Miller

## Abstract

**Background:** Research on multimorbidity—commonly defined as the presence of two or more long-term conditions—has predominantly focused on biological and clinical needs. However, unmet social care needs (SCNs), including, for example, mobility limitations, financial hardship, and social isolation, can also adversely affect health outcomes. We aimed to identify distinct clusters of individuals with multimorbidity and SCNs, and to examine their associations with 10-year all-cause mortality.

**Methods:** We analysed data from the Clinical Practice Research Datalink (CPRD) Gold and Aurum databases (Jan 1, 1987, to Dec 31, 2020) to identify adults in England living with multimorbidity. Latent class analysis was used to derive clusters based on long-term conditions and eight predefined domains of SCNs. Threshold criteria were applied to determine meaningful representation of long-term conditions and SCNs within each cluster. Associations with 10-year all-cause mortality were assessed using Cox proportional hazards models, adjusted for age, sex, ethnicity, deprivation, multimorbidity burden, SCNs, and year of multimorbidity diagnosis.

**Findings:** Amongst 7.2 million individuals with multimorbidity (mean age 54.4 years [SD 18.2]; 55% female), 37% had at least one SCN, with community care needs being the most common (28%). Four distinct clusters were identified: **Cluster 1** (mean age 37.3 years [SD 14.5]) comprised individuals with predominantly mental and behavioural conditions, no SCNs, high deprivation, and the lowest mortality risk (reference group); **Cluster 2** (mean age 57.9 years [SD 16.3]) included those with cardiovascular, musculoskeletal, and mental and behavioural conditions, no SCNs, and a higher mortality risk (adjusted hazard ratio [aHR] 1.47 [95% CI 1.45–1.48]);**Cluster 3** (mean age 62.6 years [SD 16.4]) had similar clinical profiles to cluster 2 but the highest SCN burden, lower deprivation, and the greatest mortality risk (aHR 2.83 [2.75–2.92]); **Cluster 4** (mean age 62.6 years [SD 15.2]) exhibited the highest burden of long-term conditions, two SCNs, high deprivation, and elevated mortality (aHR 1.49 [1.47– 1.51]).

**Interpretation:** We identified four distinct clusters of individuals with varying profiles of multimorbidity, social care needs, and mortality risk. These findings underscore the importance of personalised, integrated care approaches and support the development of targeted interventions to address the complex and intersecting needs of the most vulnerable populations.

**Research in Context:** *Evidence before this study:* Previous research highlights worse health outcomes for those with multimorbidities and increased social care needs. We carried out a rapid scoping review, which highlighted a lack of clarity on the definition of social care needs in this population, with no consensus in the literature. This has limited the ability to conduct research that draws meaningful comparisons across studies. To address this, we conducted a Delphi study that identified eight domains of social care needs, which we subsequently validated through a cross-sectional study. We also conducted two additional cross-sectional studies using latent class analysis of the English Longitudinal Study of Ageing (ELSA) dataset, clustering by one social care need (activities of daily living), multimorbidities, and socio-demographics.

*Added value of this study:* To our knowledge, this is the first study to apply latent class analysis to the Clinical Practice Research Datalink (CPRD) to identify clusters based on eight domains of social care needs among individuals with multimorbidity. By examining associations with 10-year all-cause mortality, we identified four distinct clusters, highlighting variations in risk and care needs. Our findings suggest that older adults (aged ≥65 years) and individuals living in the most socioeconomically deprived areas are at greatest risk. These insights offer a foundation for developing targeted, integrated care strategies tailored to the complex and overlapping social care needs and multimorbidity of vulnerable populations.

*Implications of all the available evidence:* The findings highlight the need for targeted policies that prioritise individuals aged 65 years and older with high social care needs and multimorbidity. Integrating primary and community care services, alongside expanding opportunities for social participation, may help address the complex needs of this group. In addition, tailored interventions for working-age adults—particularly those experiencing socioeconomic deprivation, from minoritised ethnic groups, or living with moderate multimorbidity but without significant social care needs—could slow health deterioration and reduce future social care demands. These insights support proactive, stratified approaches to care that respond to the intersecting challenges of multimorbidity and social care needs across the life course.

## Introduction

Multimorbidity—the coexistence of two or more chronic health conditions, including physical non-communicable diseases, mental health disorders, and long-term infectious diseases—represents a growing global health challenge. In the UK, an estimated 40% of the population is affected, with prevalence rising to 54% among those aged 65 years and older, and is projected to reach 67.8% in this age group by 2035 (1). Multimorbidity is associated with reduced quality of life, increased healthcare utilisation, and greater risk of mortality, placing substantial demands on health systems (2). The burden of multimorbidity is unequally distributed, with socioeconomically disadvantaged populations experiencing earlier onset—up to two decades sooner—than more affluent groups (1,3,4). The complexity of care is heightened by the diversity of conditions that comprise multimorbidity: younger individuals are more likely to live with conditions such as asthma, epilepsy, or autism, while older adults are more affected by cardiometabolic diseases, osteoarthritis, and depression (2). Despite this complexity, healthcare delivery remains largely focused on individual diseases, often failing to address the broader context in which these conditions coexist, including social determinants of health.

Social care needs (SCNs) are a vital but frequently overlooked dimension of multimorbidity, referring to the support required for daily living, maintaining independence, social engagement, and overall wellbeing. Approximately 50% of individuals with multimorbidity also report unmet SCNs (5), which can include mobility impairments, financial insecurity, and social isolation—factors that can further exacerbate the impact of chronic illness. Nevertheless, health and social care systems typically function in silos, contributing to fragmented and inequitable care (6–10). The interaction between multimorbidity and SCNs is of critical importance, as unmet social needs can markedly worsen health outcomes.

Emerging evidence suggests that clustering techniques may offer a more nuanced understanding of how multimorbidity and SCNs intersect, enabling the identification of distinct population subgroups defined by both health conditions and social care needs. Previous studies exploring this intersection have often been restricted to specific cohorts, such as older adults, or have considered a limited set of social care indicators (11). By incorporating eight domains of social care needs into the analytical framework (12), the present study aims to derive multimorbidity–SCN clusters and examine their association with all-cause mortality. A clearer understanding of how these domains converge across a diverse population could inform the development of integrated, person-centred care strategies to address the complex needs of individuals with multimorbidity.

## Methods

### Study design and population

This retrospective cohort study used data from the Clinical Practice Research Datalink (CPRD) GOLD and Aurum databases. We included adults aged 18 years and older with multimorbidity who were registered with a general practice (GP) in England at any time between January 1, 1987, and December 31, 2020. CPRD contains anonymised electronic health records for approximately 16 million anonymised patients, generally representative of the English population in terms of age, sex, and ethnicity (13). Individual-level data were linked across primary and secondary care records and to area-level deprivation indices, such as the Index of Multiple Deprivation (IMD).

Individuals were excluded if they were younger than 18 years at the time of multimorbidity diagnosis. Further exclusions were applied to individuals with missing follow-up time, negative follow-up duration, missing IMD information, or missing sex. Figure 1 shows the flow of patients from initial eligibility through final inclusion in the cohort.

**Figure 1:**
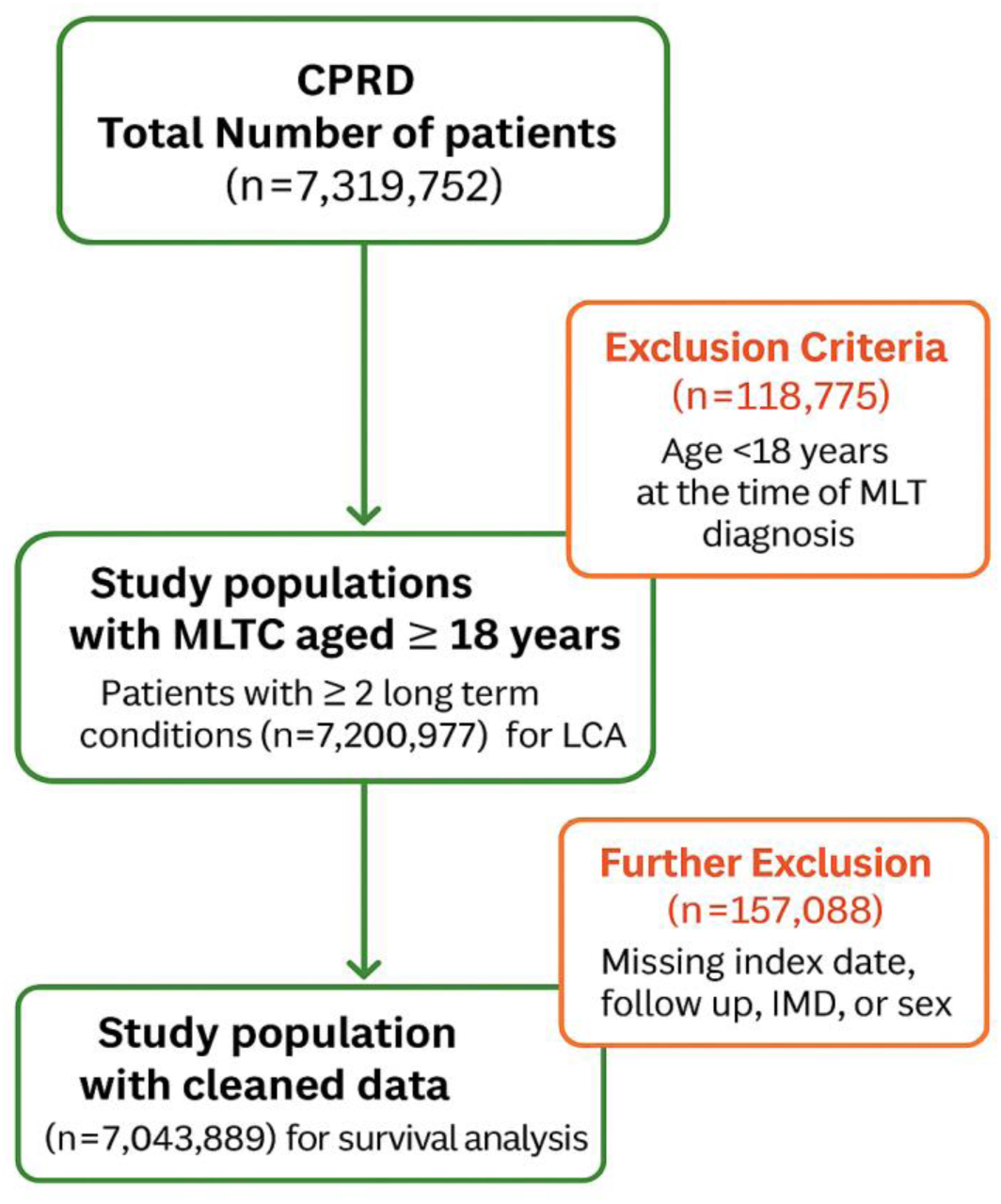
Study population flow diagram. Selection and exclusion criteria used to derive the final cohort of adults with multimorbidity from CPRD Gold and Aurum data.

### Multiple long-term conditions

Fifty-nine long-term conditions (LTCs) were identified through a national consensus study (14) (see Supplementary Table 1). From this list, the ten most prevalent LTC categories— grouped according to ICD-10 body system chapters—within the dataset were selected for use in the clustering analysis (see Figure 2). This approach is consistent with methodologies used in previous studies, balancing clinical applicability with computational efficiency (11,12). To ensure clinical relevance, the selected conditions were reviewed and validated by three practising clinicians. Multimorbidity was defined as the presence of two or more of these ten most prevalent LTCs within the dataset.

**Figure 2:**
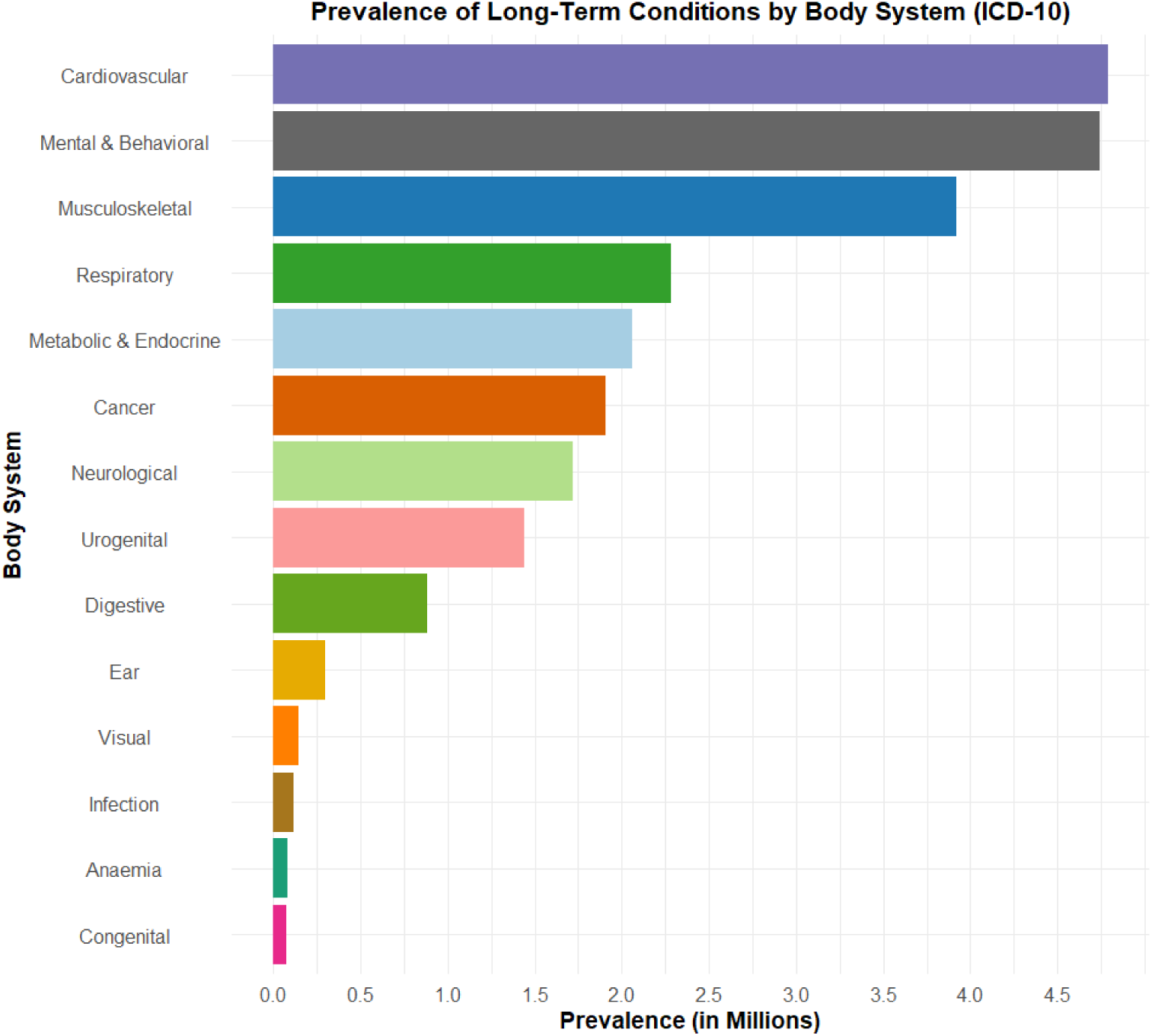
Prevalence of the ten most common long-term conditions among adults with multimorbidity in England (1987 – 2020). Data sourced from CPRD Gold and Aurum. LTC = Long-Term Conditions.

**Table 1.**
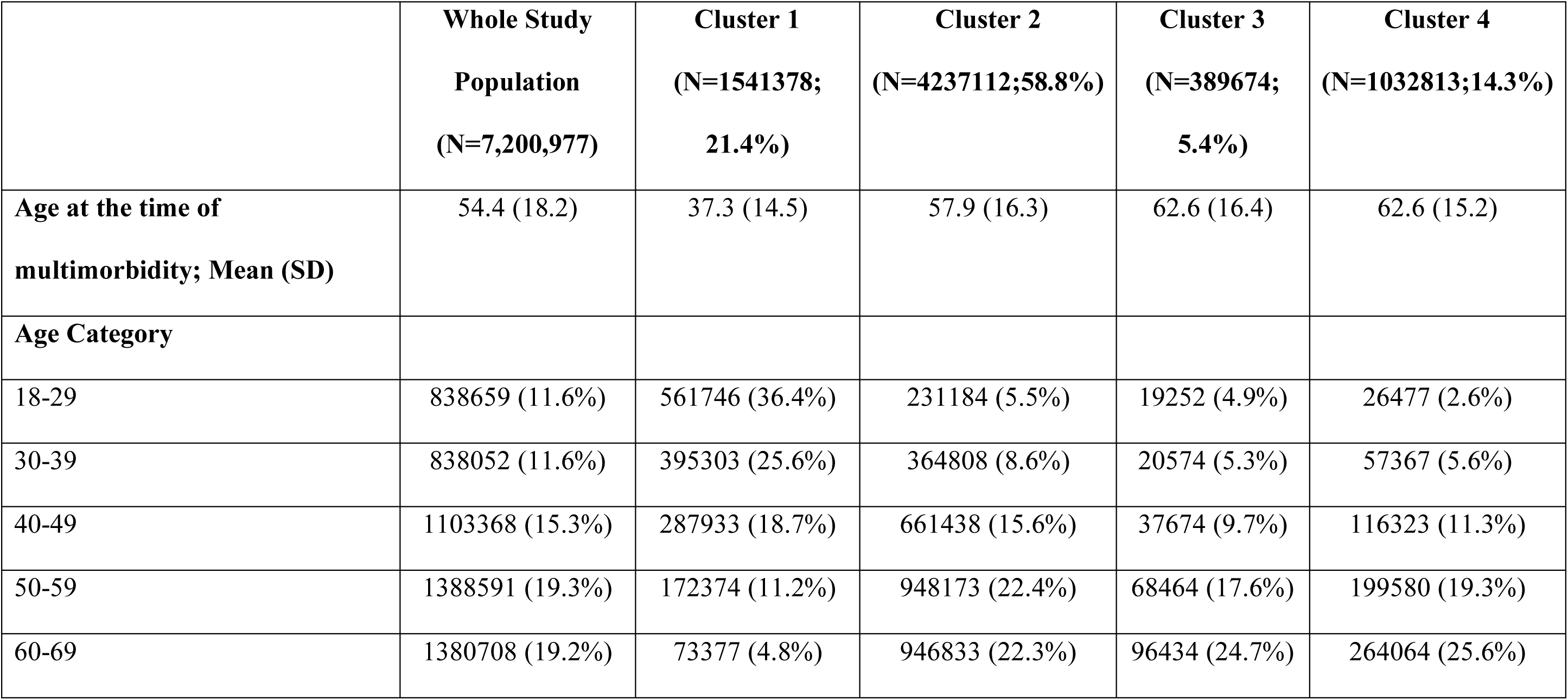
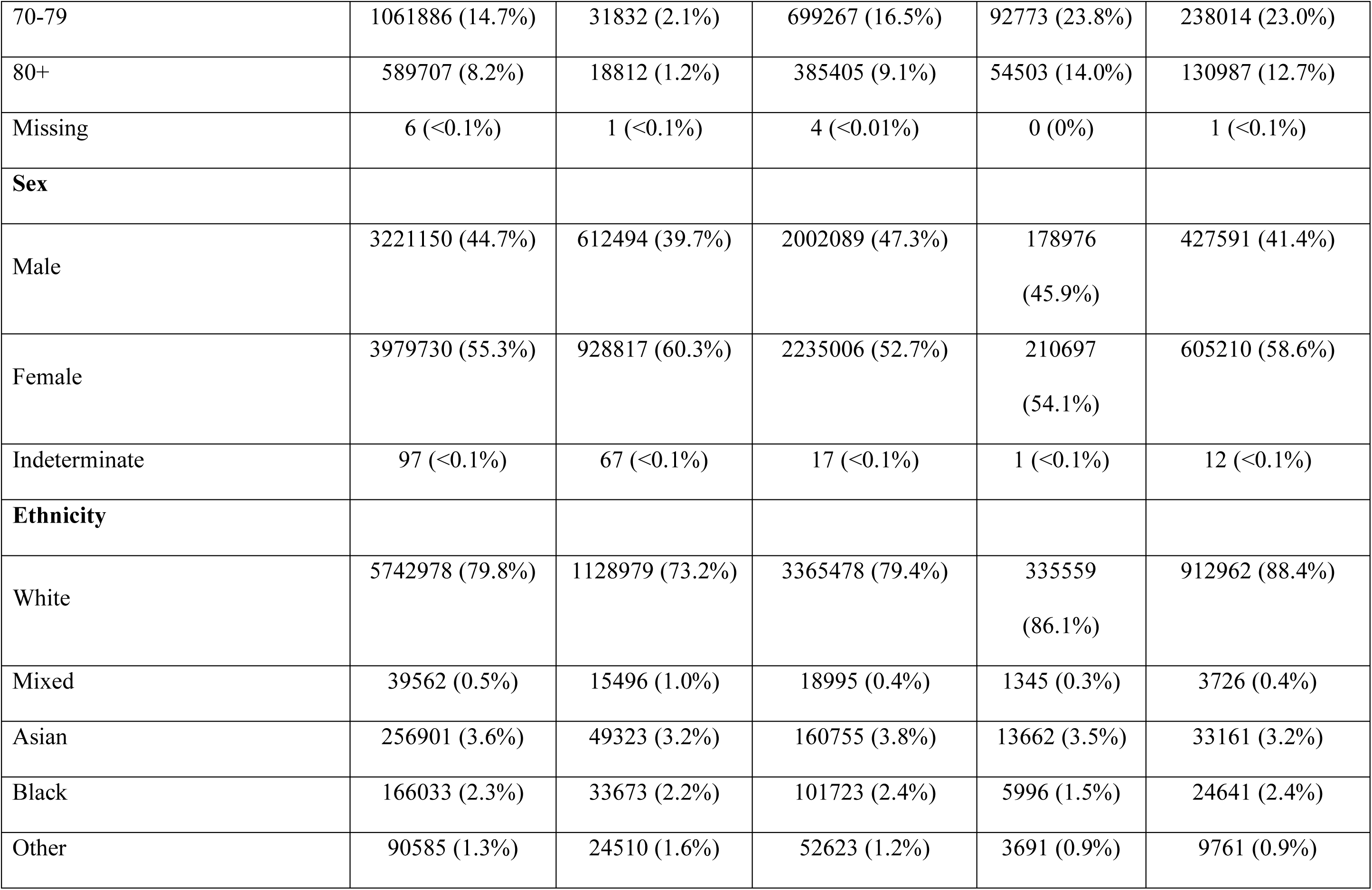
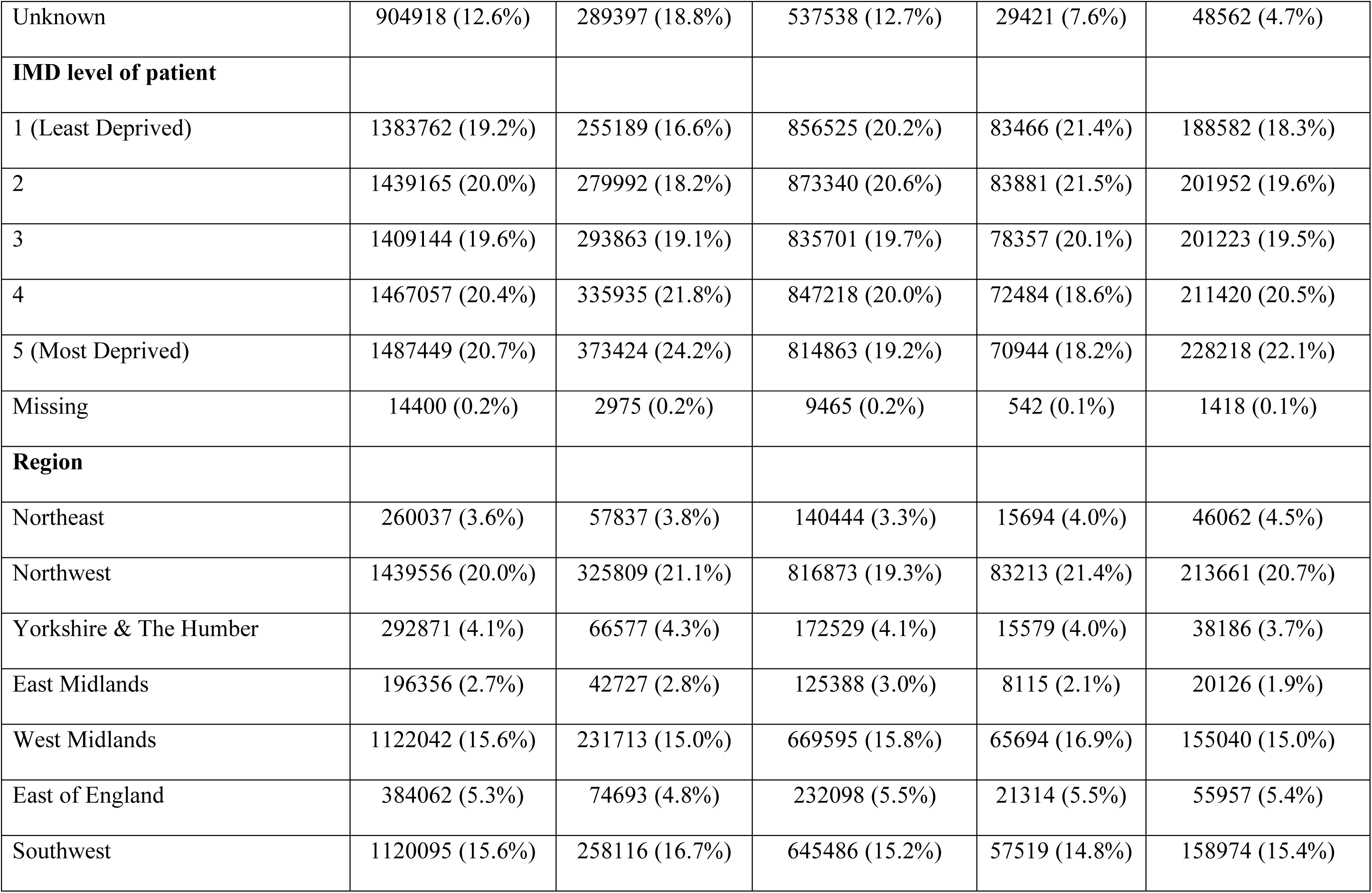
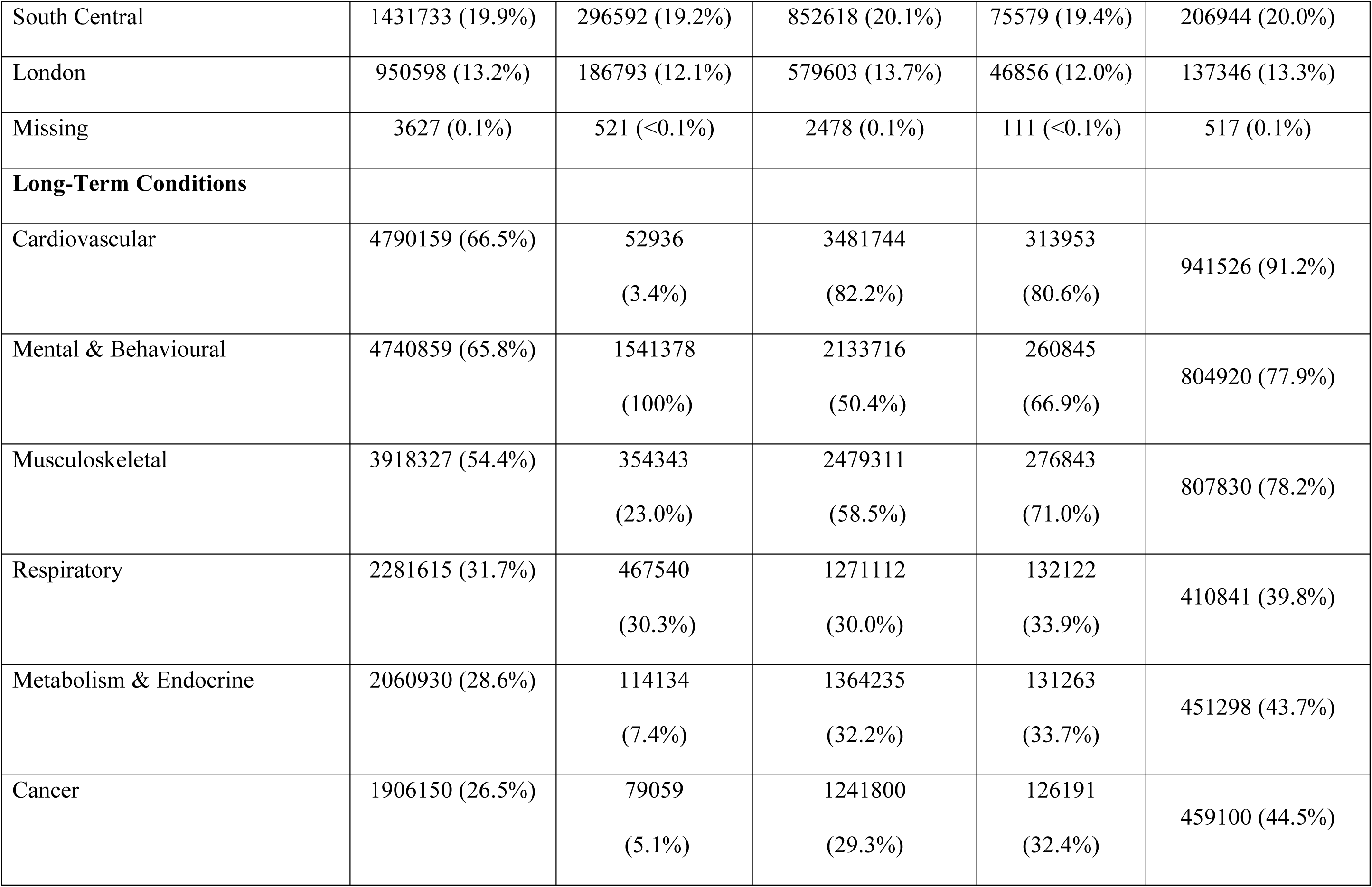
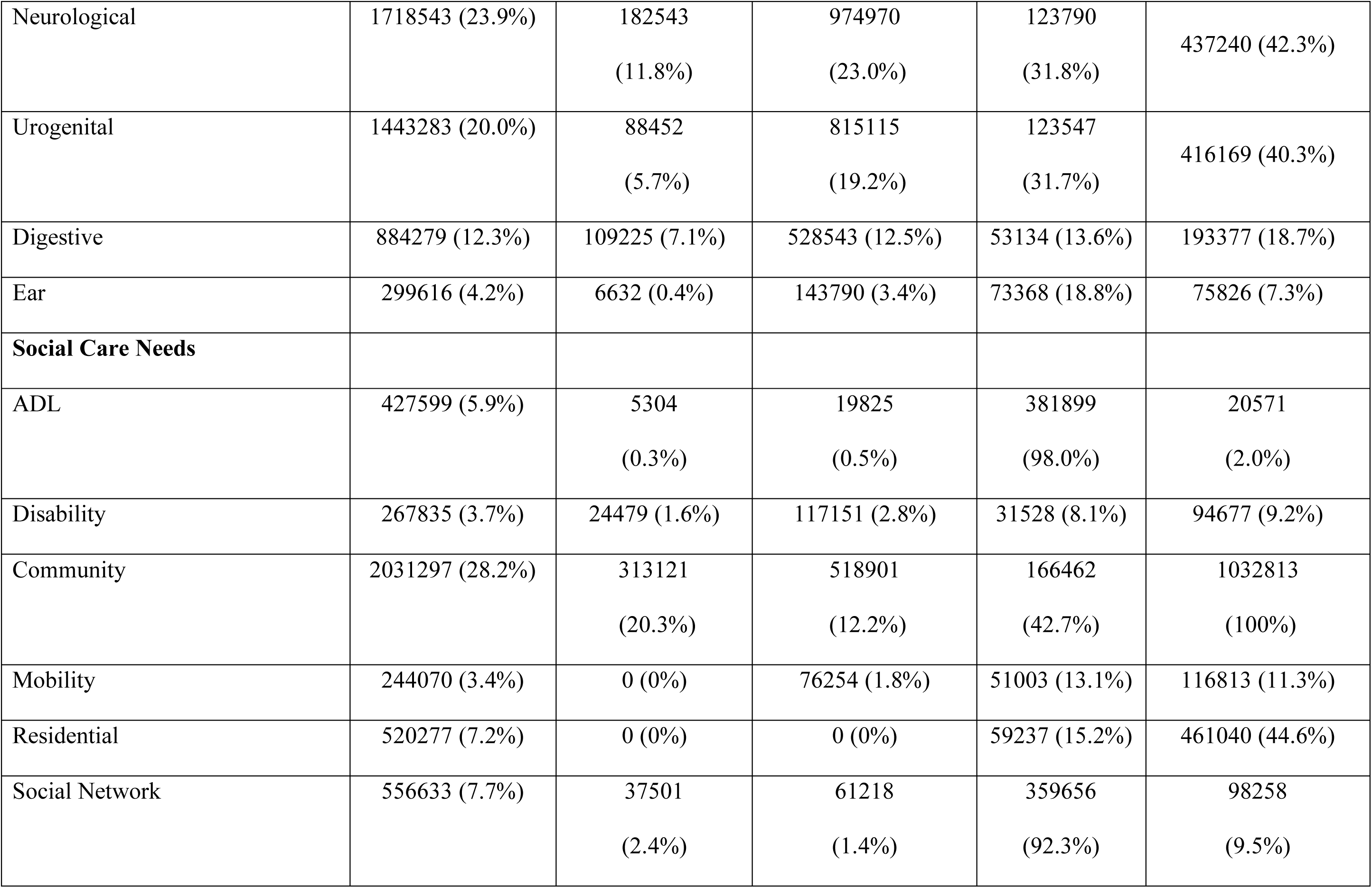
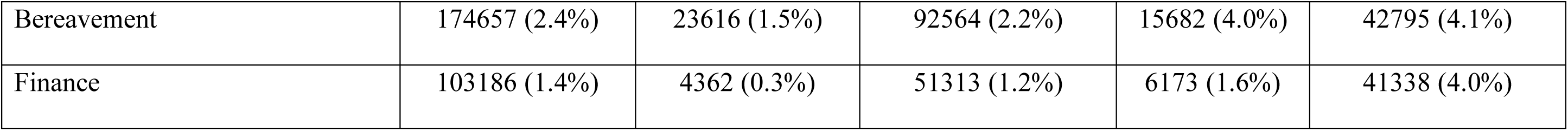
Socio-demographic characteristics, long-term conditions, and social care needs by multimorbidity clusters among 7.2 million adults in England, 1987-2020 (CPRD Gold and AURUM). Clusters were derived using latent class analysis. IMD = Index of Multiple Deprivation; LTC = Long-Term Conditions; SCN = Social Care Needs.

### Cluster model selection and classification

Model fit was assessed using the dissimilarity index (Id <0·05), classification error (close to zero), Bayesian Information Criterion (BIC), and entropy (R² close to 1), with BIC given the greatest weighting. After assessing statistical fit using these metrics and discussion with clinical experts, a four-cluster model was selected as optimal, balancing statistical performance with clinical interpretability (see Supplementary Figure 4).

Each cluster was defined by distinct combinations of LTC burden and SCNs. Figure 3 presents a heatmap of cluster-specific probabilities for LTCs and SCNs.

**Figure 3:**
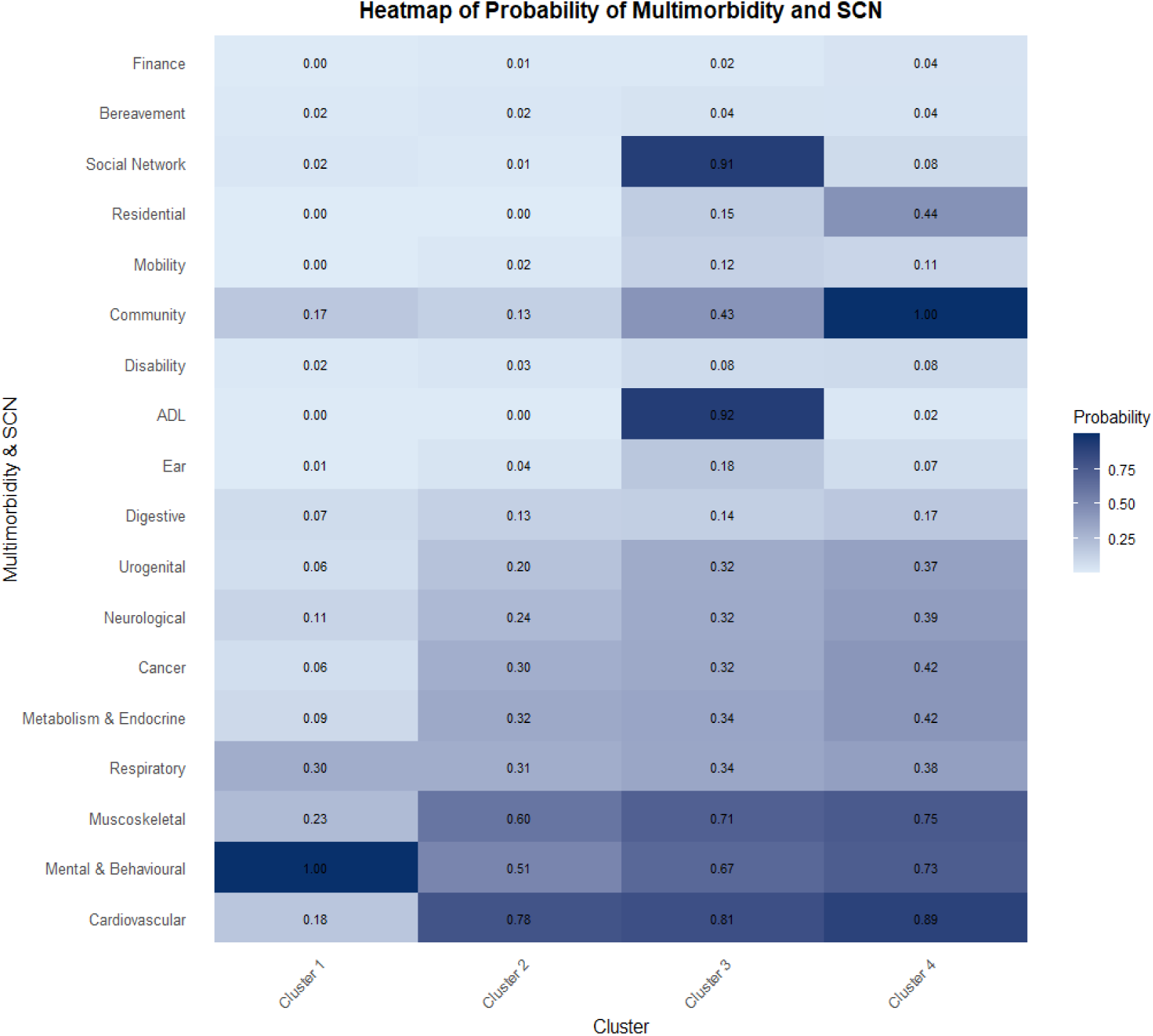
Heatmap of probability of long-term conditions and social care needs by latent cluster, CPRD 1987–2021. Probabilities of 18 features—ten long-term condition (LTC) categories and eight social care need (SCN) domains—are displayed across four clusters identified via latent class analysis using data from the Clinical Practice Research Datalink (CPRD GOLD and Aurum; N=7,200,977). Darker shading indicates a higher probability of presence.

### Cluster profiles

#### Cluster 1: Low multimorbidity, no social care needs (1 LTC, no SCN)

This cluster was characterised by the lowest burden of multimorbidity, with an almost universal presence of mental and behavioural conditions (probability 99.9%) and no social care needs. Individuals in this group had a mean age of 37.3 years (SD 14.5), making it the youngest cluster. The majority were female (60.3%), and this group had the highest proportion of individuals from non-White ethnic backgrounds. Deprivation was most prominent in this cluster, with the greatest representation in IMD quintiles 4 (21.8%) and 5 (24.2%).

#### Cluster 2: Moderate multimorbidity, no social care needs (3 LTCs, no SCN)

Cluster 2 comprised individuals with moderate multimorbidity, predominantly cardiovascular, mental and behavioural, and musculoskeletal conditions (each with probabilities >60%). The mean age was 57.9 years (SD 16.3), with the largest proportion in the 50–59-year age group (22.4%). This cluster included the highest proportion of men (47.3%) compared with the other clusters, as well as notable proportions of individuals of Asian (3.8%) and Black (2.4%) ethnicity. Deprivation was evenly distributed across IMD quintiles.

#### Cluster 3: Moderate multimorbidity, highest social care needs (3 LTCs, 3 SCNs)

Cluster 3 was distinguished by the highest burden of social care needs, particularly in relation to social network limitations, activities of daily living (ADL), and community care. The multimorbidity pattern closely resembled that of Cluster 2, with high probabilities (>67%) of cardiovascular, mental and behavioural, and musculoskeletal conditions. The mean age was 62.6 years (SD 16.4), with greater representation in the 70–79 (23.8%) and ≥80 (14.0%) age groups. Women made up 54.1% of this cluster. It was the least ethnically diverse and least deprived of all the clusters.

#### Cluster 4: High multimorbidity, moderate social care needs (5 LTCs, 2 SCNs)

This cluster exhibited the highest burden of multimorbidity, with high probabilities (>73%) of cardiovascular, mental and behavioural, musculoskeletal, metabolic/endocrine, and cancer-related conditions. Community care needs were nearly universal (99.9%), and almost half of individuals required residential support (44.0%). The mean age was 62.6 years (SD 15.2), with a predominance of individuals aged 60 years and older. Women comprised 58.6% of the cluster. It also had the second-highest proportion of individuals identifying as Black (2.4%) and a substantial representation in the most deprived IMD quintile (22.1%).

### Mortality by cluster

Kaplan-Meier curves showed clear differences in mortality risk between the four clusters. After adjusting for age, sex, ethnicity, and IMD, the mortality risk was significantly higher in all the other clusters compared to Cluster 1. The adjusted hazard ratios (aHR) for 10-year all-cause mortality were 1.47 (1.45-1.48) for Cluster 2, 2.83 (2.75 - 2.92) for Cluster 3, and 1.49 (1.47 - 1.51) for Cluster 4.

**Table 2:**
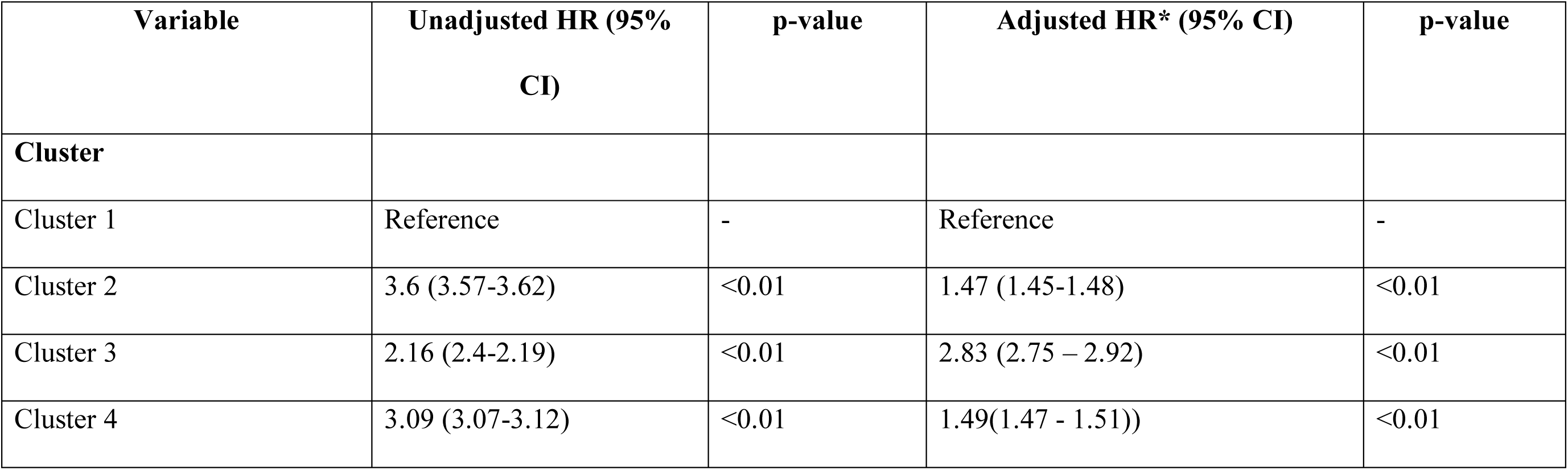
Cox proportional hazards regression results for 10-year all-cause mortality among individuals with multimorbidity, CPRD 1987–2020. Hazard ratios (HRs) and 95% confidence intervals (CIs) from unadjusted and adjusted models. Models adjusted for age category, sex, ethnicity, Index of Multiple Deprivation (IMD) quintile, and region. All p-values were <0·01. Data are from CPRD GOLD and Aurum (N=7,043,889).

## Discussion

In this population-based study of over seven million adults in England with multimorbidity, we identified four distinct clusters defined by combinations of long-term conditions (LTCs) and social care needs (SCNs) and examined their association with 10-year all-cause mortality. One in three people with multimorbidity had at least one SCN, highlighting the considerable burden on health and social care systems, and the significant impact on individuals and their carers. Among the eight SCN domains, community care, residential needs, social networking, and activities of daily living (ADL) were the most prevalent.

Mental and behavioural disorders were common across all clusters, particularly among younger, socioeconomically disadvantaged women, while physical conditions such as cardiovascular and musculoskeletal diseases predominated in older populations.

Cluster 3 exhibited the highest mortality risk, followed by clusters 4, 2, and 1. Despite lower levels of deprivation, Cluster 3 had the greatest SCN burden—particularly social network needs—which may contribute to its elevated mortality. This finding aligns with previous evidence showing that social isolation is associated with increased mortality risk, especially from cardiovascular disease, which was also common in this cluster (18). Cluster 4, which had the highest burden of LTCs and greater deprivation, had a lower prevalence of social network needs and a higher probability of receiving community care support (probability 1.0 vs 0.43 in Cluster 3), which may have had a protective effect. These results suggest that formal social care provision may mitigate the negative outcomes associated with complex multimorbidity and unmet social needs. Cluster 2 comprised individuals with moderate multimorbidity and no reported SCNs. This cluster had the highest proportion of men and people from ethnic minority groups. The absence of recorded SCNs may reflect underreporting, particularly of mental health and social needs, which are often under-recognised among men (19) and underdiagnosed in minoritised ethnic groups due to barriers to care (20). These observations highlight the limitations of routinely collected electronic health records (EHRs), which may not fully capture individual-level experiences. Further research using qualitative methods is needed to validate EHR-derived classifications and to better understand unmet need in these populations.

Our findings are consistent with existing literature indicating that cardiovascular disease is prevalent across multimorbidity clusters and contributes substantially to healthcare utilisation (21,6). Women were more likely to experience mental health conditions (22,23), which are closely linked to social deprivation (24,25). In line with previous research (16,26), the need for ADL support increased with age and was most pronounced in Cluster 3, the oldest group, which also had the highest SCN burden. The high prevalence of social network needs in this cluster likely reflects loneliness, a known risk factor for poor outcomes such as cardiovascular disease and dementia (27,28). Community care needs were especially common in clusters 3 and 4, reinforcing the intersection of older age, higher multimorbidity, and increased reliance on social support services. Given the challenges faced by carers in navigating fragmented care systems (29,30), these findings underscore the importance of coordinated service delivery.

This study has several strengths. It draws on a large, nationally representative dataset of over seven million individuals, providing substantial statistical power and generalisability. It is the first study to apply latent class analysis (LCA) to jointly classify multimorbidity and SCNs using eight comprehensive domains. Cluster identification was informed by both statistical criteria and clinical relevance, ensuring robust and interpretable groupings. Model performance was strong, with no convergence issues and adequate sample sizes across all clusters. However, some limitations should be noted. Cluster assignment in LCA is probabilistic rather than deterministic, and naming clusters may oversimplify their complexity. As an observational study, causal inference is limited, and while we adjusted for key confounders, residual confounding remains possible. Future work could incorporate time-dependent covariates to improve model accuracy. Additionally, data completeness varied across clusters, suggesting that some profiles may be more data-rich and informative than others.

These findings have important clinical and policy implications. People aged 65 years and older with high multimorbidity and SCN burden—particularly those in clusters 3 and 4— should be prioritised for integrated care interventions that combine primary, community, and social services. Addressing needs such as social isolation, mobility, and ADL support may help reduce mortality in these groups. In contrast, individuals in Cluster 2, who are of working age and show moderate multimorbidity but no recorded SCNs, may benefit from proactive identification and early support to prevent functional decline and reduce future care needs. Conditions such as mental health disorders, cardiovascular disease, and musculoskeletal problems, particularly when co-occurring with social disadvantage, require targeted prevention and management strategies to reduce health inequalities.

This is the first study to cluster individuals by both multimorbidity and social care needs, offering a more nuanced understanding of the interplay between clinical and social risk factors. The identification of distinct population subgroups with differing mortality risks provides a framework for targeted interventions and more efficient resource allocation. The clustering approach offers potential to inform national policy and commissioning decisions aimed at reducing health and social care disparities. These findings support the need for a coordinated, cross-sectoral policy response that integrates health, social care, and community systems, with a particular focus on tackling structural inequalities and ensuring that support reaches those at greatest risk.

## Contributors

AS did the statistical analysis with input from TI, NI and HDM. LS wrote the first draft of the report with input from GS, HDM, NI, TI, and AF. TI and HDM accessed and verified the data. All authors had full access to all the data in the study and HDM had final responsibility for the decision to submit for publication.

## Declaration of interests

We declare no competing interests.

## Data sharing

For this study, data were obtained from the Clinical Practice Research Datalink (CPRD) under a full license agreement, which does not permit data sharing outside the research team. However, researchers interested in replicating the study may apply directly to CPRD to gain access to the dataset.

Details of the READ and SNOMED codes used in the analysis are available on [GitHub](https://github.com/big-data-in-health/CPRD/tree/main).

## Funding

HDM has received funding from the National Institute for Health and Care Research - the Artificial Intelligence for Multiple Long-Term Conditions, or “AIM”. ’The development and validation of population clusters for integrating health and social care: A mixed-methods study on multiple long-term conditions’ (NIHR202637); receives funding from the National Institute for Health and Care Research ‘Multiple Long-Term Conditions (MLTC) Cross NIHR Collaboration (CNC)’ (NIHR207000); and receives funding from the National Institute for Health and Care Research ‘Developing and optimising an intervention prototype for addressing health and social care need in multimorbidity’ (NIHR206431). The views expressed in this publication are those of the author(s) and not necessarily those of the NHS, the National Institute for Health Research or the Department of Health and Social Care. AF was supported by the National Institute of Health Research (NIHR) Oxford Biomedical Research Centre (BRC).’

## Copyright

There are no copyright issues to declare.

## Ethical considerations

This study was approved by the Independent Scientific Advisory Committee (ISAC) for CPRD research (protocol number: 21_001667) and conducted in accordance with the relevant guidelines and regulations. CPRD data are anonymised, and individual patient consent was not required. Ethical approval was also granted by the University of Southampton Faculty of Medicine Research Committee (67953).

## SUPPLEMENTAL MATERIALS

**Supplementary Table 1:**
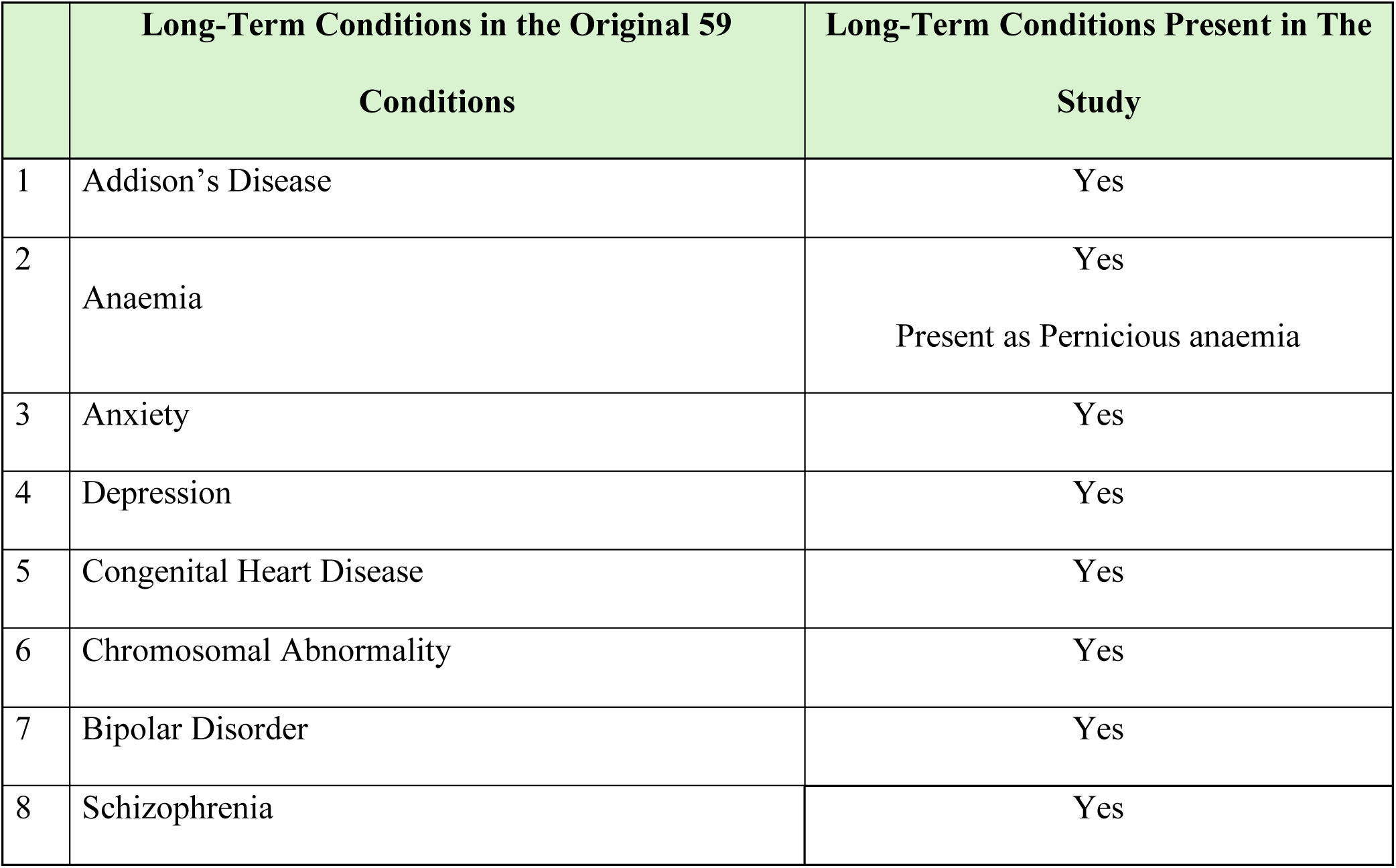
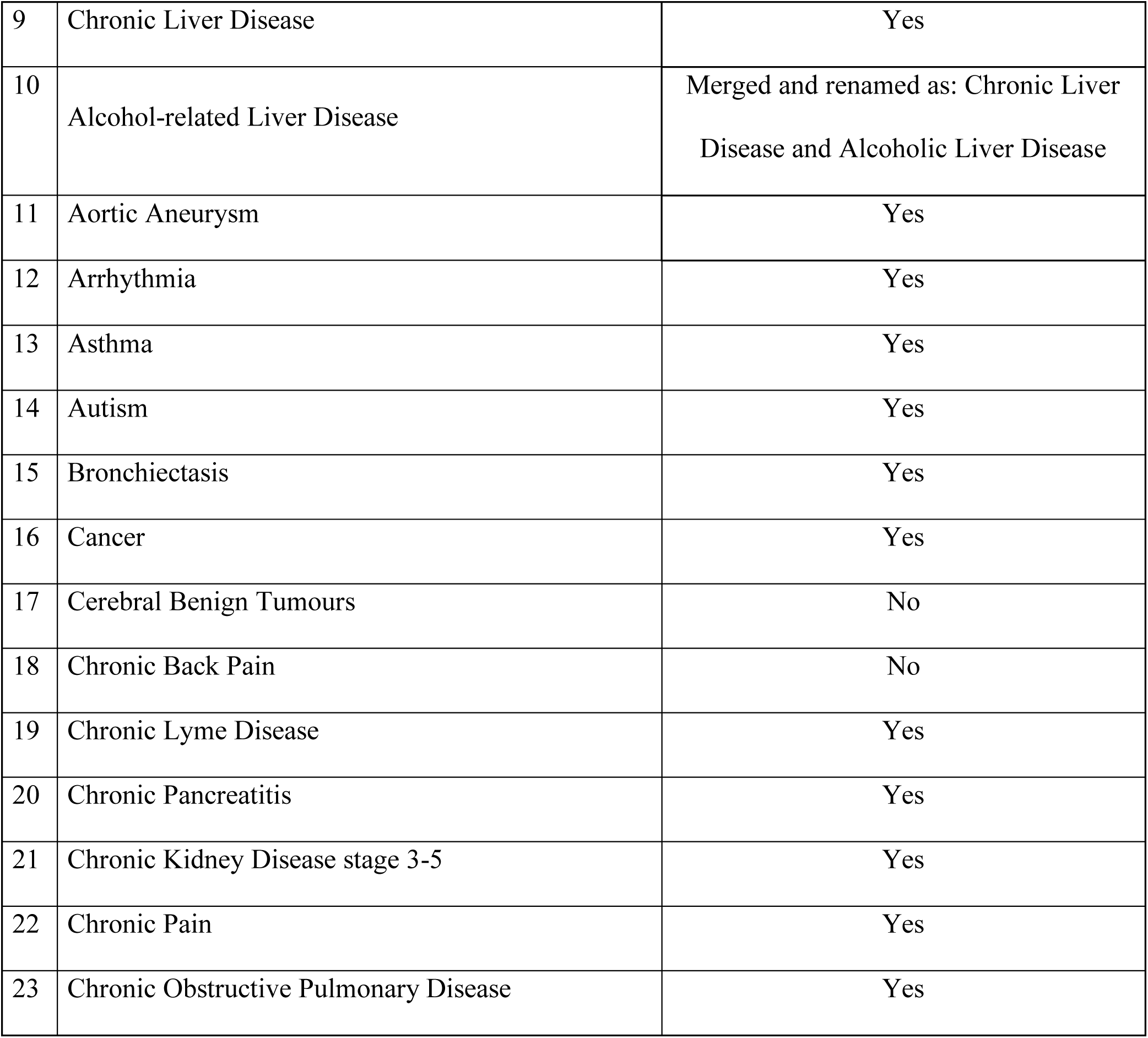
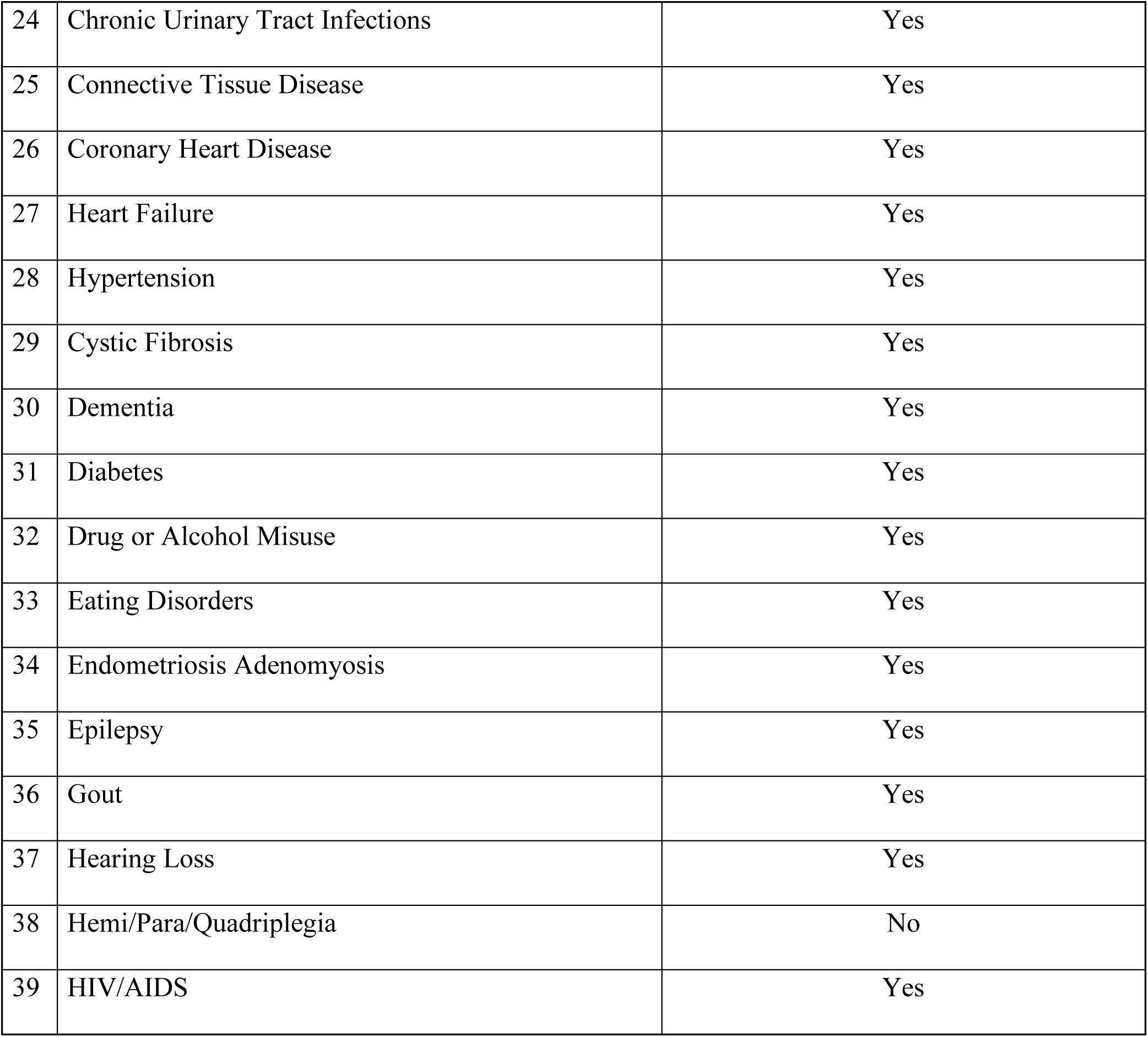
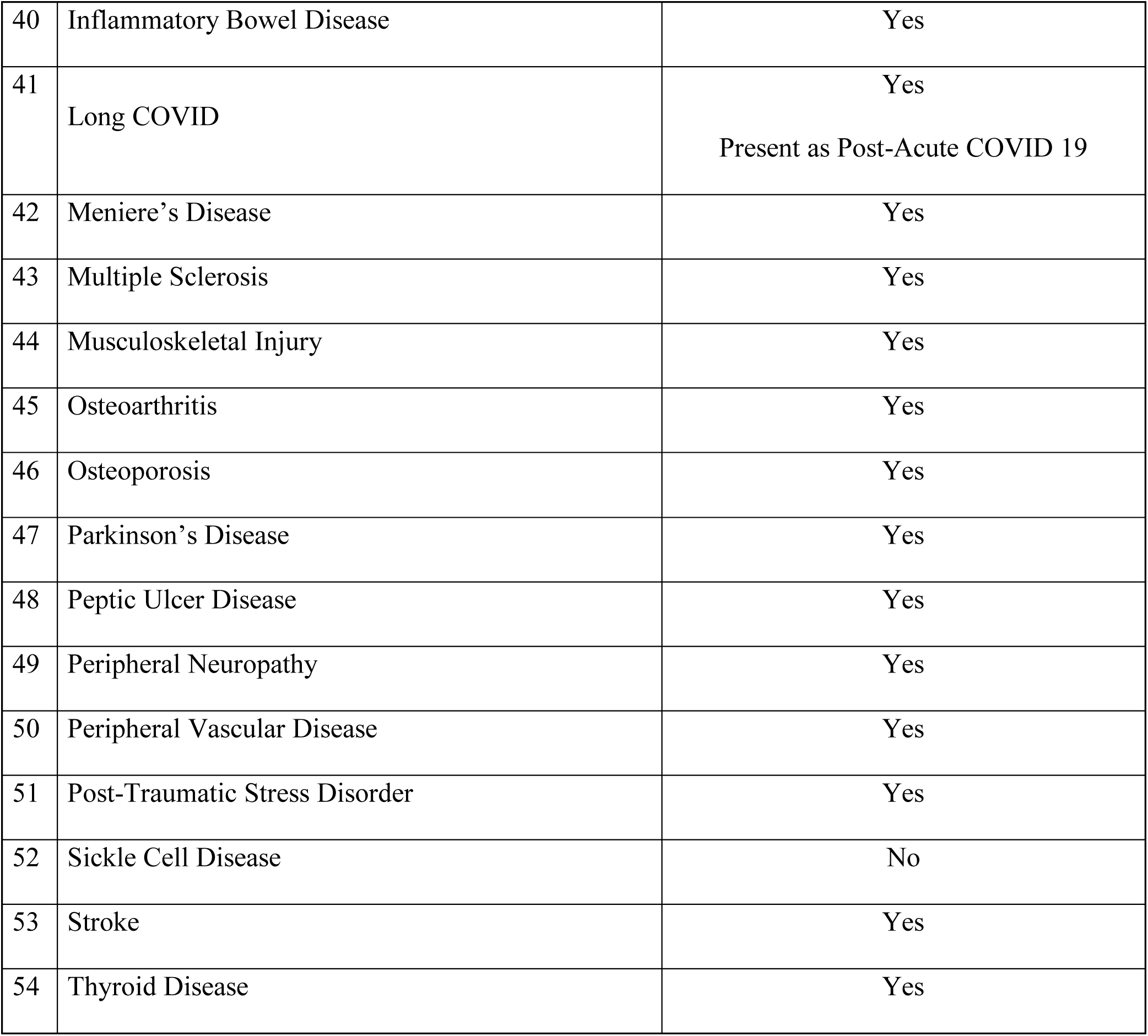
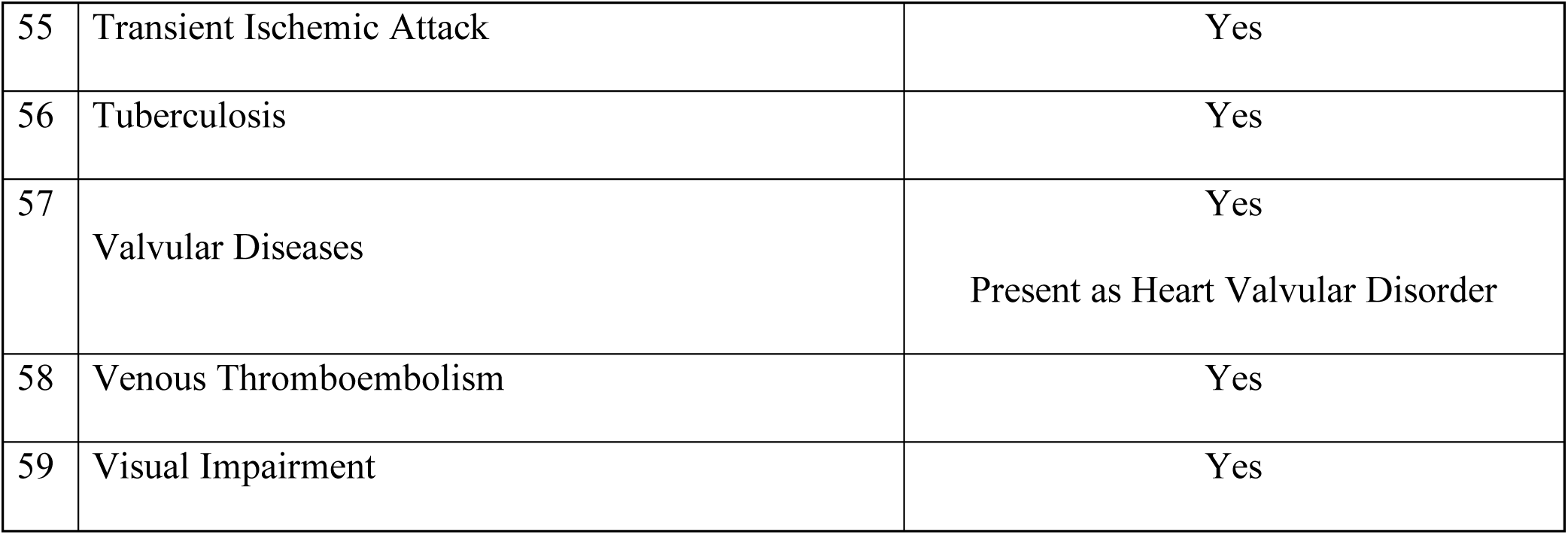
List of Long-Term Conditions (LTC) identified in the Clinical Practice Research Datalink (CPRD) Database, mapped against the original 59 LTC conditions defined through national consensus. Some conditions have been grouped based on their availability in the CPRD Database.

**Supplementary Table 2.** Characteristics of the study population aged ≥18 years with and without a record of any of the 56 long-term conditions of interest, England, 1987–2021. Data are presented as number (n) and percentage (%) unless otherwise indicated. The index date refers to the age at which the participant was diagnosed with multimorbidity. LTC = long-term condition; SD = standard deviation.

| Characteristics | CPRD Gold (n=866273) |  |  | CPRD Aurum (n=6355156) |  |  |
| --- | --- | --- | --- | --- | --- | --- |
| | None<br>3328 (0.4%) | Only 1 LTC<br>4651 (0.5%) | $\geq 2$ LTC<br>858294 (99.1%) | None<br>1 (<0.1%) | Only 1 LTC<br>12056 (0.2%) | $\geq 2$ LTC<br>6343099 (99.8%) |
| Age at index date (*mean, SD*) | NaN (NA) | NaN (NA) | 56.9 (17.8) | NaN (NA) | NaN (NA) | 54.1 (18.2) |
| Missing | 3328 (100%) | 4651 (100%) | 6 (<0.1%) | 1 (100%) | 12056 (100%) | 0 (0%) |
| Age Category |  |  |  |  |  |  |
| 18 – 29 years | 0 (0%) | 0 (0%) | 73686 (8.6%) | 0 (0%) | 0 (0%) | 765062 (12.1%) |
| 30 – 39 years | 0 (0%) | 0 (0%) | 88417 (10.3%) | 0 (0%) | 0 (0%) | 749815 (11.8%) |
| 40 – 49 years | 0 (0%) | 0 (0%) | 124465 (14.5%) | 0 (0%) | 0 (0%) | 979004 (15.4%) |
| 50 – 59 years | 0 (0%) | 0 (0%) | 164544 (19.2%) | 0 (0%) | 0 (0%) | 1224084 (19.3%) |
| 60 – 69 years | 0 (0%) | 0 (0%) | 175118 (20.4%) | 0 (0%) | 0 (0%) | 1205594 (19.0%) |
| 70 – 79 years | 0 (0%) | 0 (0%) | 145021 (16.9%) | 0 (0%) | 0 (0%) | 916867 (14.5%) |
| >80 years old | 0 (0%) | 0 (0%) | 87037 (10.1%) | 0 (0%) | 0 (0%) | 502673 (7.9%) |
| Missing | 3328 (100%) | 4651 (100%) | 6 (<0.1%) | 1 (100%) | 12056 (100%) | 0 (0%) |
| Sex |  |  |  |  |  |  |
| Male | 1413 (42.5%) | 1999 (43.0%) | 380997 (44.4%) | 0 (0%) | 5143 (42.7%) | 2840361 (44.8%) |
| Female | 1915 (57.5%) | 2652 (57.0%) | 477290 (55.6%) | 1 (100%) | 6913 (57.3%) | 3502648 (55.2%) |
| Indeterminate | 0 (0%) | 0 (0%) | 7 (<0.1%) | 0 (0%) | 0 (0%) | 90 (0.0%) |
| Index of multiple deprivation |  |  |  |  |  |  |
| 1 (least deprived) | 610 (18.3%) | 957 (20.6%) | 156519 (18.2%) | 0 (0%) | 2538 (21.1%) | 1227270 (19.3%) |
| 2 | 595 (17.9%) | 900 (19.4%) | 166993 (19.5%) | 0 (0%) | 2396 (19.9%) | 1272218 (20.1%) |
| 3 | 561 (16.9%) | 791 (17.0%) | 187984 (21.9%) | 0 (0%) | 2357 (19.6%) | 1221228 (19.3%) |
| 4 | 630 (18.9%) | 828 (17.8%) | 182021 (21.2%) | 0 (0%) | 2580 (21.4%) | 1285156 (20.3%) |
| 5 (most deprived) | 930 (27.9%) | 1168 (25.1%) | 163683 (19.1%) | 1 (100%) | 2142 (17.8%) | 1323920 (20.9%) |
| Unknown | 2 (0.1%) | 7 (0.2%) | 1094 (0.1%) | 0 (0%) | 43 (0.4%) | 13307 (0.2%) |
| Ethnicity |  |  |  |  |  |  |
| White | 2024 (60.8%) | 4014 (86.3%) | 724507 (84.4%) | 1 (100%) | 8980 (74.5%) | 5018526 (79.1%) |
| Mixed | 12 (0.4%) | 33 (0.7%) | 3215 (0.4%) | 0 (0%) | 135 (1.1%) | 36362 (0.6%) |
| Asian | 49 (1.5%) | 121 (2.6%) | 18948 (2.2%) | 0 (0%) | 730 (6.1%) | 237981 (3.8%) |
| Black | 27 (0.8%) | 83 (1.8%) | 10570 (1.2%) | 0 (0%) | 515 (4.3%) | 155649 (2.5%) |
| Other | 11 (0.3%) | 71 (1.5%) | 9537 (1.1%) | 0 (0%) | 365 (3.0%) | 81061 (1.3%) |
| Unknown | 1205 (36.2%) | 329 (7.1%) | 91517 (10.7%) | 0 (0%) | 1331 (11.0%) | 813520 (12.8%) |
| Region |  |  |  |  |  |  |
| Northeast | 0 (0%) | 59 (1.3%) | 31864 (3.7%) | 0 (0%) | 339 (2.8%) | 228182 (3.6%) |
| Northwest | 901 (27.1%) | 1144 (24.6%) | 130675 (15.2%) | 0 (0%) | 1948 (16.2%) | 1308920 (20.6%) |
| Yorkshire & The Humber | 0 (0%) | 91 (2.0%) | 53563 (6.2%) | 0 (0%) | 417 (3.5%) | 239317 (3.8%) |
| East Midlands | 675 (20.3%) | 637 (13.7%) | 39725 (4.6%) | 0 (0%) | 285 (2.4%) | 156635 (2.5%) |
| West Midlands | 0 (0%) | 124 (2.7%) | 71892 (8.4%) | 1 (100%) | 1878 (15.6%) | 1050206 (16.6%) |
| East of England | 751 (22.6%) | 845 (18.2%) | 110948 (12.9%) | 0 (0%) | 500 (4.1%) | 273133 (4.3%) |
| Southwest | 0 (0%) | 257 (5.5%) | 93961 (10.9%) | 0 (0%) | 2833 (23.5%) | 1026305 (16.2%) |
| South Central | 1001 (30.1%) | 1266 (27.2%) | 173006 (20.2%) | 0 (0%) | 2444 (20.3%) | 1258808 (19.8%) |
| London | 0 (0%) | 228 (4.9%) | 152660 (17.8%) | 0 (0%) | 1398 (11.6%) | 797966 (12.6%) |
| Missing | 0 (0%) | 0 (0%) | 0 (0%) | 0 (0%) | 14 (0.1%) | 3627 (0.1%) |

**Supplementary Table 3.** Characteristics of the study population aged ≥18 years with multimorbidity, England, 1987–2021. Data are presented as number (n) and percentage (%) unless otherwise indicated. *Anaemia includes both pernicious and sickle-cell anaemia. Indeterminate sex was excluded (n=7 in CPRD GOLD). MULTIMORBIDITY = multiple long-term conditions; SD = standard deviation.

| Characteristic | CPRD GOLD (n=858,287) |  |  | CPRD Aurum (n= 6,343,009) |  |  |
| --- | --- | --- | --- | --- | --- | --- |
|  | Total<br>n (...%) | Men<br>380997 (44.4%) | Women<br>477290 (55.6%) | Total<br>n (...%) | Men<br>2840361 (44.8%) | Women<br>3502648 (55.2%) |
| Age at time of MULTIMORBIDITY,<br>*mean (SD)* | 56.9 (17.8) | 57.5 (16.6) | 56.5 (18.8) | 54.1 (18.2) | 54.9 (17.1) | 53.4 (19.1) |
| Missing | 6 (<0.1%) | 3 (<0.1%) | 3 (<0.1%) | 0 (0%) | 0 (0%) | 0 (0%) |
| Age Category |  |  |  |  |  |  |
| 18-29 years | 73683 (8.6%) | 25678 (6.7%) | 48005 (10.1%) | 765007 (12.1%) | 273497 (9.6%) | 491510 (14.0%) |
| 30-39 years | 88415 (10.3%) | 34279 (9.0%) | 54136 (11.3%) | 749804 (11.8%) | 302165 (10.6%) | 447639 (12.8%) |
| 40-49 years | 124465 (14.5%) | 55565 (14.6%) | 68900 (14.4%) | 978994 (15.4%) | 451960 (15.9%) | 527034 (15.0%) |
| 50-59 years | 164544 (19.2%) | 80014 (21.0%) | 84530 (17.7%) | 1224075 (19.3%) | 601912 (21.2%) | 622163 (17.8%) |
| 60-69 years | 175117 (20.4%) | 87784 (23.0%) | 87333 (18.3%) | 1205593 (19.0%) | 608730 (21.4%) | 596863 (17.0%) |
| 70-79 years | 145021 (16.9%) | 65810 (17.3%) | 79211 (16.6%) | 916863 (14.5%) | 416337 (14.7%) | 500526 (14.3%) |
| ≥80 years | 87036 (10.1%) | 31864 (8.4%) | 55172 (11.6%) | 502673 (7.9%) | 185760 (6.5%) | 316913 (9.0%) |
| Missing | 6 (<0.1%) | 3 (<0.1%) | 3 (<0.1%) | 0 (0%) | 0 (0%) | 0 (0%) |
| Index of multiple deprivation |  |  |  |  |  |  |
| 1 (least deprived) | 156516 (18.2%) | 69489 (18.2%) | 87027 (18.2%) | 1227260 (19.3%) | 547259 (19.3%) | 680001 (19.4%) |
| 2 | 166991 (19.5%) | 74102 (19.4%) | 92889 (19.5%) | 1272198 (20.1%) | 568125 (20.0%) | 704073 (20.1%) |
| 3 | 187983 (21.9%) | 82804 (21.7%) | 105179 (22.0%) | 1221213 (19.3%) | 544041 (19.2%) | 677172 (19.3%) |
| 4 | 182020 (21.2%) | 80281 (21.1%) | 101739 (21.3%) | 1285133 (20.3%) | 574607 (20.2%) | 710526 (20.3%) |
| 5 (most deprived) | 163683 (19.1%) | 73815 (19.4%) | 89868 (18.8%) | 1323899 (20.9%) | 600145 (21.1%) | 723754 (20.7%) |
| Unknown | 1094 (0.1%) | 506 (0.1%) | 588 (0.1%) | 13306 (0.2%) | 6184 (0.2%) | 7122 (0.2%) |
| Ethnicity |  |  |  |  |  |  |
| White | 724503 (84.4%) | 318306 (83.5%) | 406197 (85.1%) | 5018494 (79.1%) | 2209827 (77.8%) | 2808667 (80.2%) |
| Mixed | 3215 (0.4%) | 1252 (0.3%) | 1963 (0.4%) | 36362 (0.6%) | 13711 (0.5%) | 22651 (0.6%) |
| Asian | 18948 (2.2%) | 8495 (2.2%) | 10453 (2.2%) | 237980 (3.8%) | 106056 (3.7%) | 131924 (3.8%) |
| Black | 10570 (1.2%) | 4334 (1.1%) | 6236 (1.3%) | 155648 (2.5%) | 64951 (2.3%) | 90697 (2.6%) |
| Other | 9537 (1.1%) | 4293 (1.1%) | 5244 (1.1%) | 81060 (1.3%) | 36813 (1.3%) | 44247 (1.3%) |
| Unknown | 91514 (10.7%) | 44317 (11.6%) | 47197 (9.9%) | 813465 (12.8%) | 409003 (14.4%) | 404462 (11.5%) |
| Region |  |  |  |  |  |  |
| Northeast | 31864 (3.7%) | 14240 (3.7%) | 17624 (3.7%) | 228182 (3.6%) | 103701 (3.7%) | 124481 (3.6%) |
| Northwest | 130675 (15.2%) | 59578 (15.6%) | 71097 (14.9%) | 1308894 (20.6%) | 589340 (20.7%) | 719554 (20.5%) |
| Yorkshire & The Humber | 53563 (6.2%) | 23472 (6.2%) | 30091 (6.3%) | 239315 (3.8%) | 108119 (3.8%) | 131196 (3.7%) |
| East Midlands | 39725 (4.6%) | 17500 (4.6%) | 22225 (4.7%) | 156635 (2.5%) | 70209 (2.5%) | 86426 (2.5%) |
| West Midlands | 71892 (8.4%) | 31835 (8.4%) | 40057 (8.4%) | 1050192 (16.6%) | 474745 (16.7%) | 575447 (16.4%) |
| East of England | 110947 (12.9%) | 49168 (12.9%) | 61779 (12.9%) | 273131 (4.3%) | 121583 (4.3%) | 151548 (4.3%) |
| Southwest | 93960 (10.9%) | 41256 (10.8%) | 52704 (11.0%) | 1026291 (16.2%) | 455526 (16.0%) | 570765 (16.3%) |
| South Central | 173002 (20.2%) | 76153 (20.0%) | 96849 (20.3%) | 1258789 (19.8%) | 555683 (19.6%) | 703106 (20.1%) |
| London | 152659 (17.8%) | 67795 (17.8%) | 84864 (17.8%) | 797953 (12.6%) | 359607 (12.7%) | 438346 (12.5%) |
| Missing | 0 (0%) | 0 (0%) | 0 (0%) | 3627 (0.1%) | 1848 (0.1%) | 1779 (0.1%) |
| **Multiple long-term conditions** |  |  |  |  |  |  |
| Addison's disease | 1,478 (0.2%) | 592 (0.2%) | 886 (0.2%) | 11,541 (0.2%) | 4689 (0.2%) | 6852 (0.2%) |
| Anaemia * |  | 3378 (0.9%) | 6621 (1.4%) | 70,475 (1.1%) | 23965 (0.8%) | 46510 (1.3%) |
| Aneurysm | 21,484 (2.5%) | 15507 (4.1%) | 5977 (1.3%) | 140,578 (2.2%) | 102486 (3.6%) | 38092 (1.1%) |
| Anxiety | 237,900 (27.7%) | 82648 (21.7%) | 155252 (32.5%) | 2,085,653<br>(32.9%) | 741305 (26.1%) | 1344348 (38.4%) |
| Arrhythmia | 240,682 (28.0%) | 121959 (32.0%) | 118723 (24.9%) | 1,985,163<br>(31.3%) | 999910 (35.2%) | 985253 (28.1%) |
| Asthma | 200,002 (23.3%) | 79444 (20.9%) | 120558 (25.3%) | 1,636,133<br>(25.8%) | 664169 (23.4%) | 971964 (27.7%) |
| Autism | 2,287 (0.3%) | 1514 (0.4%) | 773 (0.2%) | 23,807 (0.4%) | 15746 (0.6%) | 8061 (0.2%) |
| Bipolar disorder | 11,576 (1.3%) | 4446 (1.2%) | 7130 (1.5%) | 89,215 (1.4%) | 34986 (1.2%) | 54229 (1.5%) |
| Bronchiectasis | 8,446 (1.0%) | 3705 (1.0%) | 4741 (1.0%) | 81,292 (1.3%) | 36394 (1.3%) | 44898 (1.3%) |
| Chromosomal abnormality | 1,166 (0.1%) | 568 (0.1%) | 598 (0.1%) | 7,699 (0.1%) | 3529 (0.1%) | 4170 (0.1%) |
| Chronic kidney disease | 129,755 (15.1%) | 54945 (14.4%) | 74810 (15.7%) | 908,478 (14.3%) | 392180 (13.8%) | 516298 (14.7%) |
| Chronic liver disease | 27,215 (3.2%) | 16685 (4.4%) | 10530 (2.2%) | 203,110 (3.2%) | 126121 (4.4%) | 76989 (2.2%) |
| Chronic Lyme disease | 323 (<0.1%) | 152 (0.0%) | 171 (0.0%) | 4,372 (0.1%) | 1875 (0.1%) | 2497 (0.1%) |
| Chronic obstructive pulmonary disease | 135,360 (15.8%) | 70323 (18.5%) | 65037 (13.6%) | 879,562 (13.9%) | 459554 (16.2%) | 420008 (12.0%) |
| Chronic pancreatic disease | 5,369 (0.6%) | 3408 (0.9%) | 1961 (0.4%) | 37,027 (0.6%) | 23335 (0.8%) | 13692 (0.4%) |
| Chronic primary pain | 24,604 (2.9%) | 9510 (2.5%) | 15094 (3.2%) | 234,907 (3.7%) | 90144 (3.2%) | 144763 (4.1%) |
| Chronic urinary tract infection | 41,104 (4.8%) | 7311 (1.9%) | 33793 (7.1%) | 289,655 (4.6%) | 55087 (1.9%) | 234568 (6.7%) |
| Congenital heart disease | 7,456 (0.9%) | 3651 (1.0%) | 3805 (0.8%) | 57,053 (0.9%) | 27593 (1.0%) | 29460 (0.8%) |
| Connective tissue disease | 82,231 (9.6%) | 26343 (6.9%) | 55888 (11.7%) | 544,232 (8.6%) | 175330 (6.2%) | 368902 (10.5%) |
| Coronary heart disease | 227,776 (26.5%) | 126137 (33.1%) | 101639 (21.3%) | 1,437,334<br>(22.7%) | 808510 (28.5%) | 628824 (18.0%) |
| Cystic fibrosis | 623 (0.1%) | 159 (0.0%) | 464 (0.1%) | 2,802 (<0.1%) | 854 (<0.1%) | 1948 (0.1%) |
| Dementia | 69,475 (8.1%) | 26797 (7.0%) | 42678 (8.9%) | 497,195 (7.8%) | 194809 (6.9%) | 302386 (8.6%) |
| Depression | 335,624 (39.1%) | 121732 (32.0%) | 213892 (44.8%) | 2,436,361<br>(38.4%) | 901910 (31.8%) | 1534451 (43.8%) |
| Diabetes mellitus | 165,873 (19.3%) | 88582 (23.3%) | 77291 (16.2%) | 1,206,134<br>(19.0%) | 648670 (22.8%) | 557464 (15.9%) |
| Drug or alcohol misuse | 238,255 (27.8%) | 136966 (35.9%) | 101289 (21.2%) | 1,950,706<br>(30.8%) | 1105641 (38.9%) | 845065 (24.1%) |
| Eating disorder | 7,168 (0.8%) | 751 (0.2%) | 6417 (1.3%) | 62,287 (1.0%) | 6165 (0.2%) | 56122 (1.6%) |
| Endometriosis | 17,362 (2.0%) | 13 (0.0%) | 17349 (3.6%) | 145,216 (2.3%) | 160 (0.0%) | 145056 (4.1%) |
| Epilepsy | 32,440 (3.8%) | 15711 (4.1%) | 16729 (3.5%) | 225,916 (3.6%) | 111608 (3.9%) | 114308 (3.3%) |
| Gout | 69,467 (8.1%) | 50239 (13.2%) | 19228 (4.0%) | 488,468 (7.7%) | 359863 (12.7%) | 128605 (3.7%) |
| Haematological cancer | 236,804 (27.6%) | 115908 (30.4%) | 120896 (25.3%) | 1,668,427<br>(26.3%) | 811970 (28.6%) | 856457 (24.5%) |
| Hearing impairment that cannot be corrected | 21,466 (2.5%) | 10522 (2.8%) | 10944 (2.3%) | 220,564 (3.5%) | 106167 (3.7%) | 114397 (3.3%) |
| Heart failure | 142,044 (16.5%) | 71565 (18.8%) | 70479 (14.8%) | 885,143 (14.0%) | 453071 (16.0%) | 432072 (12.3%) |
| Heart valve disorder | 77,318 (9.0%) | 38039 (10.0%) | 39279 (8.2%) | 549,315 (8.7%) | 271354 (9.6%) | 277961 (7.9%) |
| HIV/AIDS | 1,280 (0.1%) | 924 (0.2%) | 356 (0.1%) | 15,497 (0.2%) | 10969 (0.4%) | 4528 (0.1%) |
| Hypertension | 359,819 (41.9%) | 166698 (43.8%) | 193121 (40.5%) | 2,770,340<br>(43.7%) | 1309895 (46.1%) | 1460445 (41.7%) |
| Inflammatory bowel disease | 17,155 (2.0%) | 7693 (2.0%) | 9462 (2.0%) | 122,407 (1.9%) | 56480 (2.0%) | 65927 (1.9%) |
| Long-term musculoskeletal problem due to injury | 141,772 (16.5%) | 46398 (12.2%) | 95374 (20.0%) | 1,092,136 (17.2%) | 384467 (13.5%) | 707669 (20.2%) |
| Meniere's disease | 8,162 (1.0%) | 2827 (0.7%) | 5335 (1.1%) | 55,875 (0.9%) | 19557 (0.7%) | 36318 (1.0%) |
| Metastatic cancer | 79,633 (9.3%) | 38971 (10.2%) | 40662 (8.5%) | 497,244 (7.8%) | 245351 (8.6%) | 251893 (7.2%) |
| Multiple sclerosis | 3,881 (0.5%) | 1109 (0.3%) | 2772 (0.6%) | 28,687 (0.5%) | 8452 (0.3%) | 20235 (0.6%) |
| Osteoarthritis | 331,727 (38.6%) | 132575 (34.8%) | 199152 (41.7%) | 2,341,660 (36.9%) | 953173 (33.6%) | 1388487 (39.6%) |
| Osteoporosis | 87,891 (10.2%) | 16700 (4.4%) | 71191 (14.9%) | 641,029 (10.1%) | 124038 (4.4%) | 516991 (14.8%) |
| Paralysis | 27,360 (3.2%) | 13424 (3.5%) | 13936 (2.9%) | 174,370 (2.7%) | 87374 (3.1%) | 86996 (2.5%) |
| Parkinson's disease | 11,244 (1.3%) | 6407 (1.7%) | 4837 (1.0%) | 80,628 (1.3%) | 47108 (1.7%) | 33520 (1.0%) |
| Peptic ulcer | 76,410 (8.9%) | 42528 (11.2%) | 33882 (7.1%) | 481,477 (7.6%) | 272362 (9.6%) | 209115 (6.0%) |
| Peripheral neuropathy | 92,761 (10.8%) | 35697 (9.4%) | 57064 (12.0%) | 680,821 (10.7%) | 270269 (9.5%) | 410552 (11.7%) |
| Peripheral vascular disease | 72,249 (8.4%) | 37760 (9.9%) | 34489 (7.2%) | 414,298 (6.5%) | 219876 (7.7%) | 194422 (5.6%) |
| Post-acute COVID-19 | 44 (<0.1%) | 16 (0.0%) | 28 (0.0%) | 17,016 (0.3%) | 5654 (0.2%) | 11362 (0.3%) |
| Post-traumatic stress disorder | 4,008 (0.5%) | 1935 (0.5%) | 2073 (0.4%) | 80,153 (1.3%) | 37146 (1.3%) | 43007 (1.2%) |
| Schizophrenia | 10,594 (1.2%) | 5767 (1.5%) | 4827 (1.0%) | 90,531 (1.4%) | 49164 (1.7%) | 41367 (1.2%) |
| Stroke | 106,408 (12.4%) | 51473 (13.5%) | 54935 (11.5%) | 701,584 (11.1%) | 346113 (12.2%) | 355471 (10.1%) |
| Thyroid disease | 110,362 (12.9%) | 24726 (6.5%) | 85636 (17.9%) | 765,040 (12.1%) | 171218 (6.0%) | 593822 (17.0%) |
| Transient ischaemic attack | 53,025 (6.2%) | 25552 (6.7%) | 27473 (5.8%) | 335,590 (5.3%) | 162591 (5.7%) | 172999 (4.9%) |
| Tuberculosis | 8,695 (1.0%) | 4110 (1.1%) | 4585 (1.0%) | 68,749 (1.1%) | 33896 (1.2%) | 34853 (1.0%) |
| Venous thromboembolic disease | 63,321 (7.4%) | 27850 (7.3%) | 35471 (7.4%) | 429,370 (6.8%) | 191890 (6.8%) | 237480 (6.8%) |
| Visual impairment that cannot be corrected | 17,291 (2.0%) | 6699 (1.8%) | 10592 (2.2%) | 126,598 (2.0%) | 50650 (1.8%) | 75948 (2.2%) |
| **Social care variables** |  |  |  |  |  |  |
| Activities of daily living needs (ADL) | 48,163 (5.6%) | 22626 (5.9%) | 25537 (5.4%) | 379,443 (6.0%) | 175120 (6.2%) | 204323 (5.8%) |
| Disability needs | 18,550 (2.2%) | 8597 (2.3%) | 9953 (2.1%) | 249,287 (3.9%) | 113719 (4.0%) | 135568 (3.9%) |
| Community care needs | 217,750 (25.4%) | 90374 (23.7%) | 127376 (26.7%) | 1,813,523<br>(28.6%) | 782565 (27.6%) | 1030958 (29.4%) |
| Mobility needs | 36,672 (4.3%) | 14423 (3.8%) | 22249 (4.7%) | 207,398 (3.3%) | 82964 (2.9%) | 124434 (3.6%) |
| Residential status needs | 47,806 (5.6%) | 18115 (4.8%) | 29691 (6.2%) | 472,468 (7.4%) | 179831 (6.3%) | 292637 (8.4%) |
| Social network needs | 23,912 (2.8%) | 9477 (2.5%) | 14435 (3.0%) | 532,726 (8.4%) | 234667 (8.3%) | 298059 (8.5%) |
| Bereavement needs | 29,267 (3.4%) | 8391 (2.2%) | 20876 (4.4%) | 145,388 (2.3%) | 41383 (1.5%) | 104005 (3.0%) |
| Financial support needs | 33,943 (4.0%) | 16420 (4.3%) | 17523 (3.7%) | 69,244 (1.1%) | 31306 (1.1%) | 37938 (1.1%) |

**Supplementary Table 4.** Summary statistics for three- to six-cluster models derived from latent class analysis, CPRD 1987–2021. Model performance indicators are shown for latent class solutions specifying three to six clusters. LL = log likelihood; NP = number of parameters; DF = degrees of freedom; AIC = Akaike Information Criterion; BIC = Bayesian Information Criterion; CE = classification error; Id = dissimilarity index. Lower AIC, BIC, and CE values reflect better model fit; dissimilarity index values <0.05 indicate good separation between clusters. Data are based on adults aged ≥18 years with multimorbidity from the CPRD GOLD and Aurum datasets.

| Number of<br>Clusters | LL | NP | DF | AIC | BIC | CE | Id | Entropy<br>R<sup>2</sup> |
| --- | --- | --- | --- | --- | --- | --- | --- | --- |
| 3 | -49611114 | 56 | 262087 | 99222340 | 99223112 | 0.1533 | 0.0438 | 0.6336 |
| 4 | -48992651 | 75 | 262068 | 97985452 | 97986486 | 0.1367 | 0.0115 | 0.7238 |
| 5 | -48764580 | 94 | 262049 | 97529349 | 97530645 | 0.1403 | 0.0124 | 0.7322 |
| 6 | -48655419 | 113 | 262030 | 97311065 | 97312623 | 0.2129 | 0.0310 | 0.6515 |

**Supplemental Table 5:** Item response probabilities for each long-term condition and social care need domain by latent cluster, CPRD 1987–2021.

| **Item Probabilities** | **Cluster 1** | **Cluster 2** | **Cluster 3** | **Cluster 4** |
| --- | --- | --- | --- | --- |
| Cardiovascular | 0.18 | 0.78 | 0.81 | 0.89 |
| Mental & Behavioural | 0.99 | 0.51 | 0.67 | 0.73 |
| Musculoskeletal | 0.23 | 0.60 | 0.71 | 0.75 |
| Respiratory | 0.30 | 0.31 | 0.34 | 0.38 |
| Metabolism &<br>Endocrine | 0.09 | 0.32 | 0.34 | 0.42 |
| Cancer | 0.06 | 0.30 | 0.32 | 0.42 |
| Neurological | 0.11 | 0.24 | 0.32 | 0.39 |
| Urogenital | 0.06 | 0.20 | 0.32 | 0.37 |
| Digestive | 0.07 | 0.13 | 0.14 | 0.17 |
| Ear | 0.01 | 0.04 | 0.18 | 0.07 |
| ADL | <0.01 | <0.01 | 0.92 | 0.02 |
| Disability | 0.02 | 0.03 | 0.08 | 0.08 |
| Community | 0.17 | 0.13 | 0.43 | 0.99 |
| Mobility | <0.01 | 0.02 | 0.12 | 0.11 |
| Residential | <0.01 | <0.01 | 0.15 | 0.44 |
| Social Network | <0.01 | 0.01 | 0.91 | 0.08 |
| Bereavement | <0.01 | 0.02 | 0.04 | 0.04 |
| Finance | <0.01 | 0.01 | 0.02 | 0.04 |

**Supplementary Table 6:** Variables included in each of the eight social care needs (SCN) domains. This table outlines the specific variables mapped to each of the eight SCN domains derived through ontological and clinical consensus processes. Domains include: (1) activities of daily living, (2) disability, (3) community care, (4) mobility, (5) residential status, (6) social network, (7) bereavement, and (8) financial support. These domains were used to identify social care needs among adults with multimorbidity in the CPRD GOLD and Aurum datasets.

| Social Care Needs | Components |
| --- | --- |
| ADL | • Received professional care or support for:<br>Activities of Daily Living (ADL)<br>• Difficulty handling food<br>• Assistance with toiletry<br>• Support with gardening<br>• Help with taking a shower<br>• Challenges using stairs<br>• Assistance with physical activities<br>• Help with bathing<br>• Difficulty using appliances<br>• Support with travelling<br>• Personal care assistance<br>• Managing medication<br>• Help performing clerical activities<br>• Assistance with cleaning the home<br>• Difficulty in walking |
|  | • Support with performing intellectual activities<br>• Help using non-verbal and verbal communication<br>• Assistance with hearing<br>• Support understanding verbal and written language<br>• Difficulty speaking |
| Disability | • Has mental disability<br>• Has learning disability<br>• Disability related to speech<br>• Sensory disability<br>• Disability quantified by the expanded disability status scale<br>• Receiving disability living allowance<br>• Relative has disability<br>• Disability assessed by the OPCS dexterity disability scale<br>• Severity of disability<br>• Physical disability<br>• On social care disability register<br>• Assessed for disability<br>• Disability assessed by the OPCS continence disability scale<br>• Multiple and complex disability |
|  | • Disability related to hearing |
| Community Care | • Seen by community occupational therapy<br>• Seen by social service<br>• Seen by community drug team<br>• Seen by community nurse<br>• Seen by community physiotherapy<br>• Seen by community audiologist<br>• Seen by community specialist<br>• Seen by community midwife<br>• Seen by community podiatrist<br>• Seen by community matron<br>• Seen by community care navigator<br>• Seen by community dietician<br>• Admitted to community hospital<br>• Seen by community mental health team<br>• Seen by community pharmacy<br>• Referral/appointment for DMARD (Disease-modifying anti-rheumatic drug)<br>• Referral to community doctor |
| Mobility | • Able to mobilise using mobility aid<br>• Waterlow scale assessing risk of developing pressure sore(s)<br>• Difficulty with mobility |
|  | • Do not mobilise using aid<br>• Difficulty mobilising indoors<br>• Mobility assessed using the Rivermead Mobility Index<br>• Difficulty managing stairs<br>• Mobilise using aid<br>• Mobility assessed using the modified elderly mobility scale<br>• Mobility assessed using the shuttle walking test<br>• Difficulty mobilising outdoors<br>• Difficulty walking |
| Residential Status | • Lives in care home<br>• Lives in hospice<br>• Lives in nursing home<br>• Lives in own home |
| Social Network | • Difficulty with communication<br>• Support provided professionally to assist social networking and social engagement<br>• Referral to social prescriber<br>• Difficulty travelling to take part in social network activities<br>• Difficulty with social participation |
| Bereavement | • Death of sibling<br>• Death of child |
|  | • Death of partner<br>• Death of parent<br>• Death of a family member |
| Financial | • Receiving disability living allowance<br>• Received mobility allowance<br>• Difficulty managing personal finance<br>• Support for housing |

**Supplementary Figure 1.**
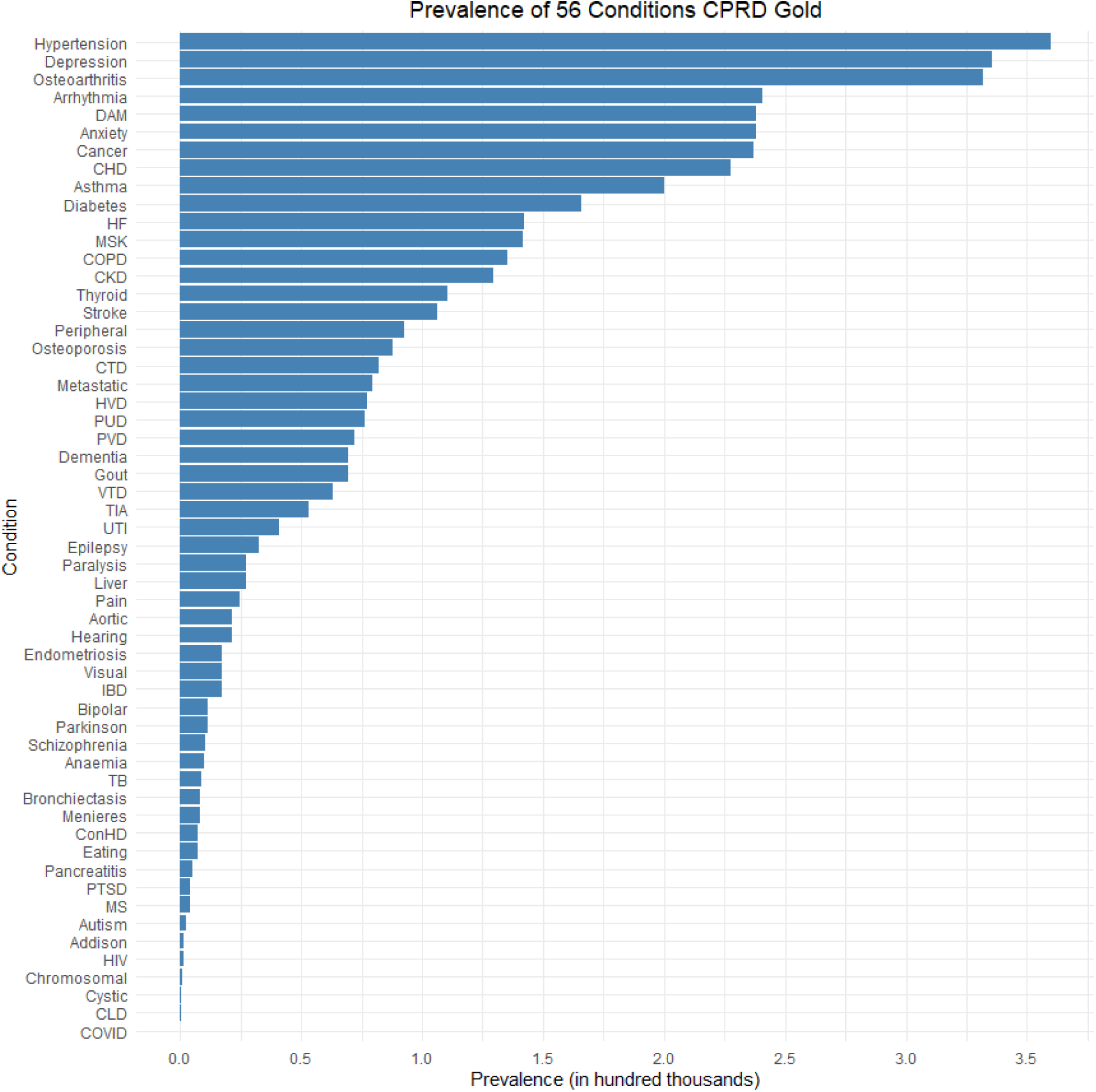
The prevalence of each 56 long-term conditions from CPRD Gold.

**Supplementary Figure 2.**
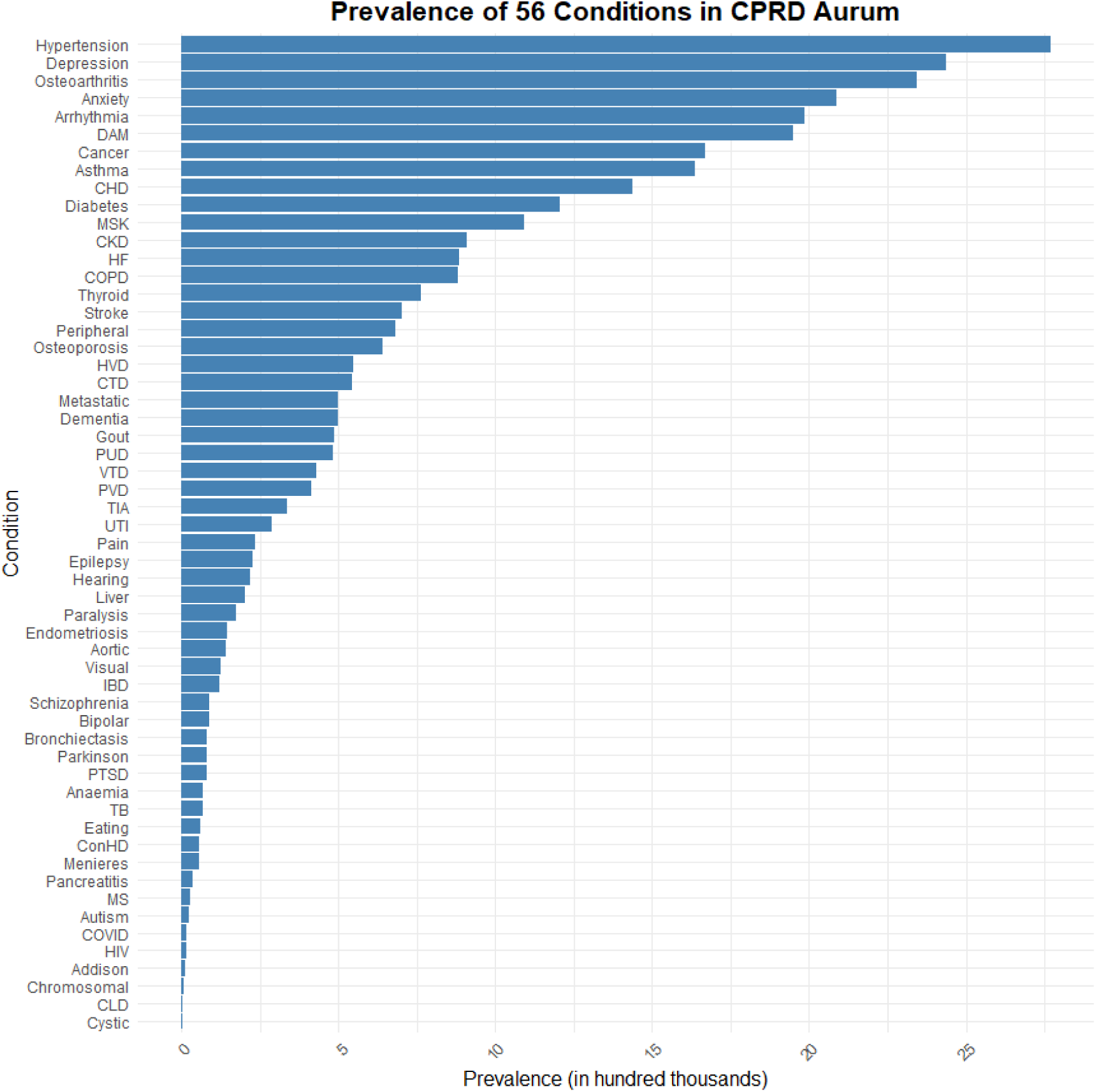
The prevalence of each 56 long-term conditions from CPRD Aurum.

**Supplementary Figure 3.**
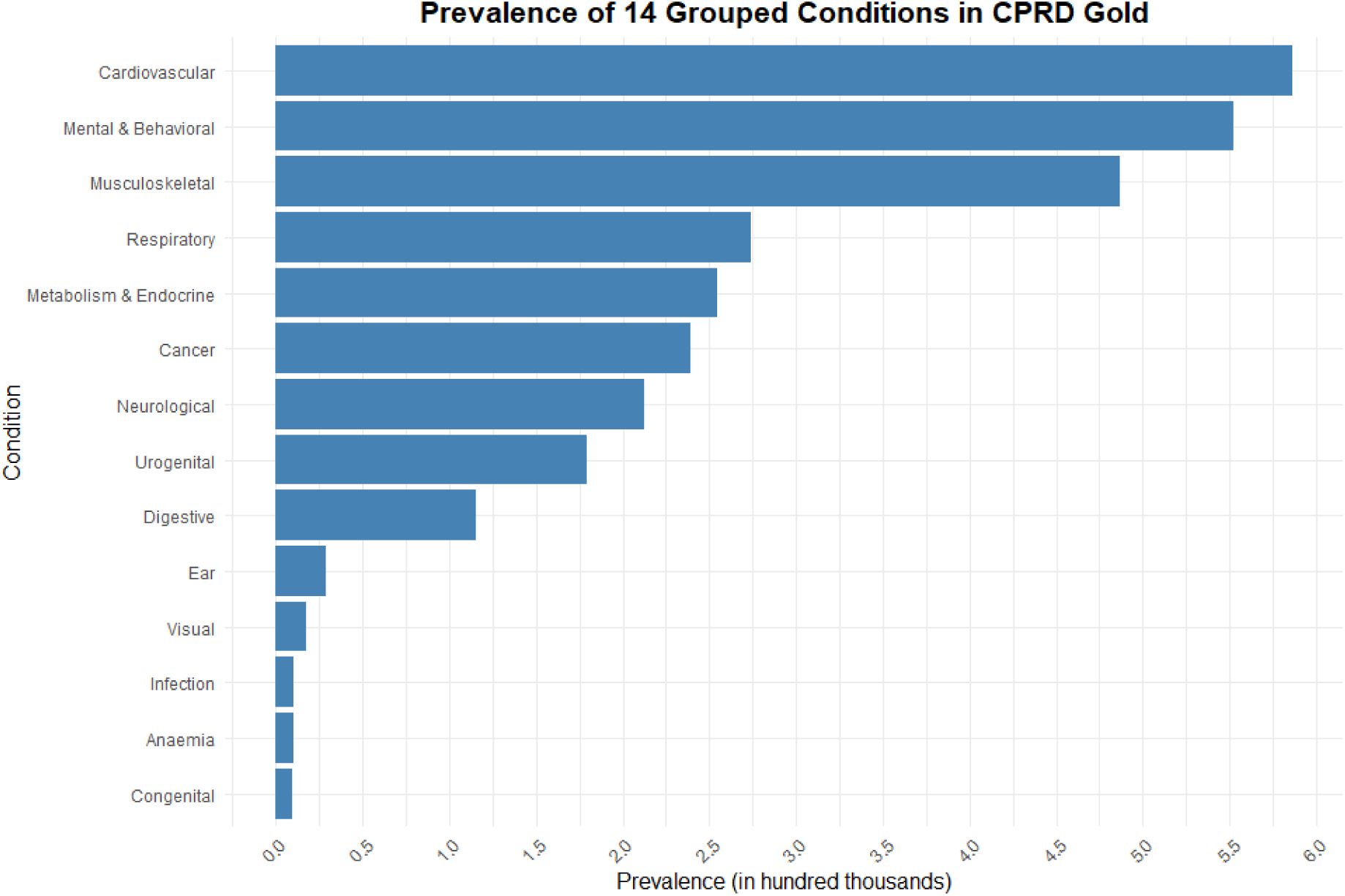
The prevalence of each 14 grouped long-term conditions from CPRD Gold.

**Supplementary Figure 4.**
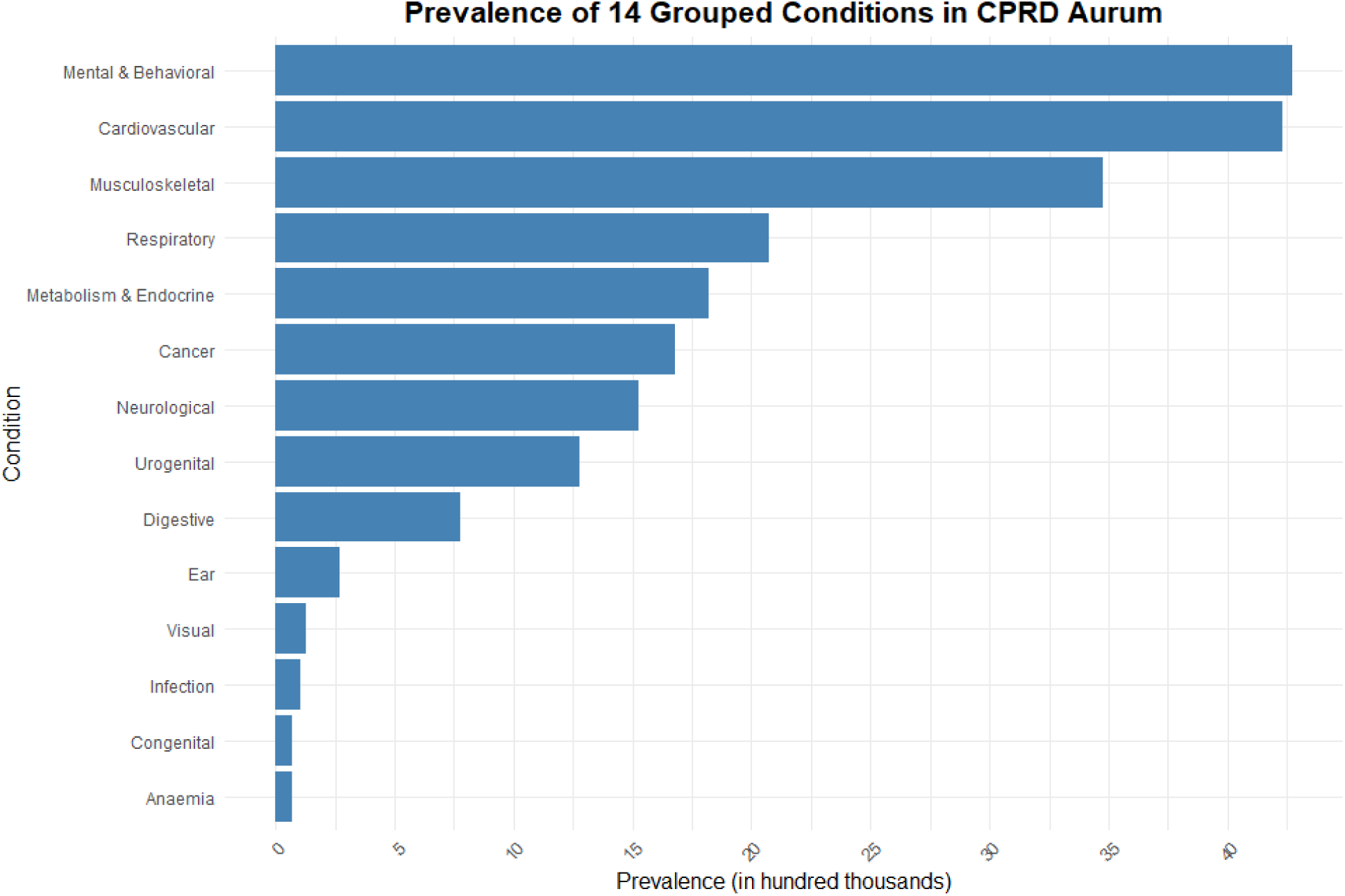
The prevalence of each 14 grouped long-term conditions from CPRD Aurum.

**Supplemental Figure 5.**
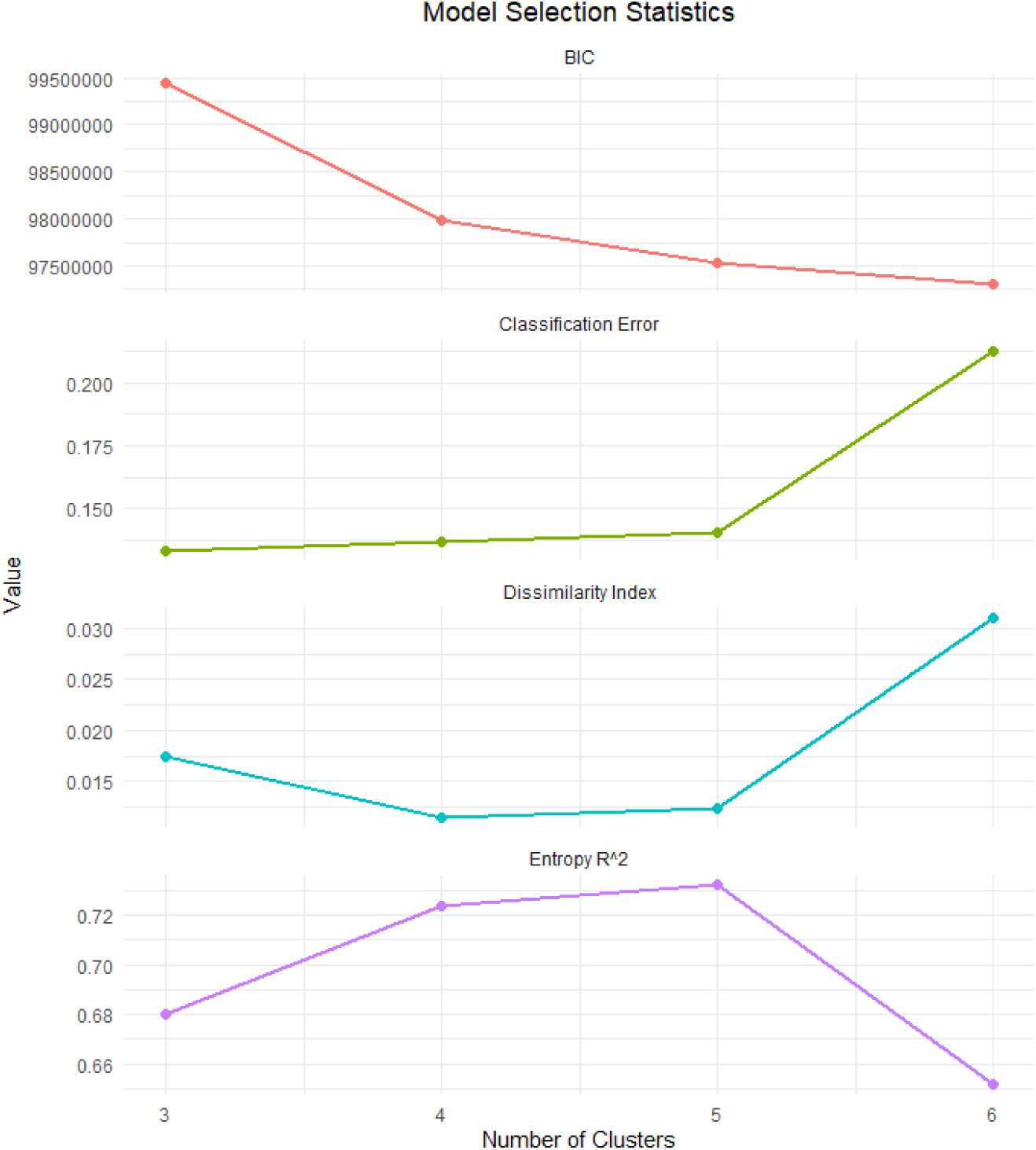
Model fit statistics for latent class models with 3 to 6 clusters. Dissimilarity Index, Classification Error, Bayesian Information Criterion (BIC), and entropy (R²) are shown for each model. The four-cluster solution was selected based on the overall pattern of fit statistics and clinical interpretability.

## Notes

### Competing Interest Statement

The authors have declared no competing interest.

